# Risk of selection and timelines for the continued spread of artemisinin and partner drug resistance in Africa

**DOI:** 10.1101/2024.08.28.24312699

**Authors:** Oliver J. Watson, Salome Muchiri, Abby Ward, Cecile Meier-Sherling, Victor Asua, Thomas Katairo, Tom Brewer, Gina Cuomo-Dannenburg, Peter Winskill, Jeffrey A Bailey, Lucy Okell, Graziella Scudu, Aaron M. Woolsey

## Abstract

**long:** The introduction of artemisinin combination therapies (ACTs) has significantly reduced the burden of *Plasmodium falciparum* malaria, yet the emergence of artemisinin partial resistance (ART-R) as well as partner drug resistance threatens these gains. Recent confirmations of prevalent *de novo* ART-R mutations in Africa, in particular in Rwanda, Uganda and Ethiopia, underscore the urgency of addressing this issue in Africa. Our objective is to characterise this evolving resistance landscape in Africa and understand the speed with which ART-R will continue to spread. We produce estimates of both ART-R and partner drug resistance by bringing together WHO, WWARN and MalariaGen Pf7k data on antimalarial resistance in combination with a literature review. We integrate these estimates within a mathematical modelling approach, aincorporating to estimate parameters known to impact the selection of ART-R for each malaria-endemic country and explore scenarios of ART-R spread and establishment. We identify 16 malaria-endemic countries in Africa to prioritise for surveillance and future deployment of alternative antimalarial strategies, based on ART-R reaching greater than 10% prevalence by 2040 under current malaria burden and effective-treatment coverage. If resistance continues to spread at current rates with no change in drug policy, we predict that partner drug resistance will emerge and the mean percentage of treatment failure across Africa will reach 30.74% by 2060 (parameter uncertainty range: 24.98% - 34.54%). This translates to an alarming number of treatment failures, with 52,980,600 absolute cases of treatment failure predicted in 2060 in Africa (parameter uncertainty range: 26,374,200 - 93,672,400) based on current effective treatment coverage. Our results provide a refined and updated prediction model for the emergence of ART-R to help guide antimalarial policy and prioritise future surveillance efforts and innovation in Africa. These results put into stark context the speed with which antimalarial resistance may spread in Africa if left unchecked, confirming the need for swift and decisive action in formulating antimalarial treatment policies focused on furthering malaria control and containing antimalarial resistance in Africa.

**short:** The rise of artemisinin partial resistance (ART-R) and increasing partner drug tolerance by *Plasmodium falciparum* malaria in Africa threatens to undo malaria control efforts. Recent confirmations of de novo ART-R markers in Rwanda, Uganda, and Ethiopia highlight the urgent need to address this threat in Africa, where the vast majority of cases and deaths occur. This study characterises the resistance landscape and predicts the spread of antimalarial resistance across Africa. We estimate and map the current levels of resistance markers related to artesmisinin and its partner drugs using WHO, WWARN, and MalariaGen Pf7k data. We combine these estimates with current malaria transmission and treatment data and use an established individual-based model of malaria resistance to simulate future resistance spread. We identify 16 African countries at highest risk of ART-R for prioritisation of enhanced surveillance and alternative antimalarial strategies. We project that, without policy changes, ART-R will exceed 10% in these regions by 2040. By 2060, if resistance spreads unchecked, we predict mean treatment failure rates will reach 30.74% (parameter uncertainty range: 24.98% - 34.54%) across Africa. This alarming spread of resistance is predicted to cause 52.98 million treatment failures (uncertainty range: 26.37 million - 93.67 million) in 2060. The impact of antimalarial resistance in Africa, if left unchecked, would hugely damage efforts to reduce malaria burden. Our results underscore the critical need for swift policy action to contain resistance and guide future surveillance and intervention efforts.

## Introduction

The introduction of artemisinin combination therapies (ACTs) as global first-line treatments for uncomplicated *Plasmodium falciparum* malaria has been vital in reducing malaria burden globally and slowing the emergence of artemisinin partial resistance (ART-R). However, ART-R has emerged and threatens to reverse recent gains in malaria control. Initially identified in Cambodia in the early 2000s, ART-R has spread extensively in Southeast Asia (SE Asia), with validated mutations in *pfkelch13* responsible for ART-R now published by the WHO.^2^ Recently, ART-R has emerged de novo in Africa, where the vast majority of cases and deaths occur, with validated *pfkelch13* mutations identified at highest prevalence in Rwanda^3^ and Uganda^4^. This development has prompted calls for an improved response to the emerging threat in Africa,^5^ including increasing molecular surveillance for ART-R, changing treatment strategies to protect the therapeutic lifespan of artemisinin, and more widespread and rapid assessment of ACT therapeutic efficacy.^6^ In response to the growing threat of ART-R, the World Health Organization (WHO) published a strategy in 2022 to respond to antimalarial drug resistance in Africa.^7^

The spread of antimalarial resistance in SE Asia led to significant ACT treatment failures, with 35-45% of patients treated with ACT failing therapy in many regions due to parasites with both artemisinin and partner drug resistance.^8^ The emergence of artemisinin resistance in Africa has raised concerns that similar trends may unfold in Africa. However, the spread of resistance may not mirror the trajectory seen in SE Asia due to higher transmission intensities in Africa, which is believed to both increase competition between wild-type and resistant parasites as well as contribute to higher recombination rates that cause multiple resistance mutations to disassociate more quickly.^9^ Further, parasites with *pfkelch13* mutations have not yet been documented to cause >10% 28-day treatment failure rates in therapeutic efficacy studies (TES) of artemether-lumefantrine (AL) in the current hotspots of ART-R emergence in Rwanda and Uganda.^5,10^ However, growing evidence to suggests that treatment failure is increasing in Africa. The observation of decreasing AL efficacy in TES conducted in travellers returning from the continent (Halsey and Plucinski 2023) and the frequency of incomplete AL drug adherence in Africa^13^ suggest that AL efficacy in routine settings may be lower than observed in trial settings.^14^ In clinical trials where all doses of AL are not supervised, 28-day PCR corrected treatment failure rates for AL have been observed >10% even in settings without any *pfkelch13* mutations,^15^ suggesting that failure in ART-R areas may be higher. Patterns of directional selection in genetic markers associated with increased lumefantrine tolerance have been observed after AL treatment.^16^ Further, observational studies in Uganda reveal fast increases in ART-R frequency comparable to those observed during the initial years of artemisinin resistance emergence in SE Asia.^17^ Together, these suggest treatment failures in AL-treated patients occur and may increase due to rising lumefantrine tolerance and ART-R.

Mathematical models of malaria and resistance play a crucial role in understanding and managing the spread of antimalarial drug resistance, particularly under the frameworks proposed by the WHO’s “Strategy to respond to antimalarial drug resistance in Africa.”^7^ These models help identify the risk factors driving the emergence and spread of resistance, such as drug pressure, within-host competition and human behaviour, which all influence the evolution and spread of resistant malaria strains.^18^ By simulating various scenarios and interventions, these models help in assess the impact of drug resistance on malaria control and eradication efforts, providing a robust platform for decision-making.^19^ Modelling also supports specific missions of the WHO resistance strategy, notably Pillar 4, which emphasises understanding and tracking resistance. It aids in mapping the risk of antimalarial resistance,^20^ highlighting regions and populations where resistance is more likely to develop and spread, which enables surveillance and interventions to be targeted effectively.^21^ Furthermore, models can forecast the impact of changes in diagnostic practices and treatment strategies in reducing drug pressure, thus informing the optimization of these tools to delay or prevent the emergence of resistance.^6^ Integrating detailed genetic and epidemiological data into these models can refine predictions and enhance the design of tailored strategies to manage and mitigate resistance.^22^

In this study, we incorporate recent advances in our understanding of the current frequency of antimalarial resistance and construct geostatistical models of the frequency of antimalarial resistance in Africa. Using current estimates and trends in antimalarial resistance, we project the continued spread of resistance in sub-Saharan Africa over the next 40 years using a previously published individual-based model of malaria transmission to model the selection of antimalarial resistance.^19^ These maps can be used to guide ongoing surveillance efforts and where the future deployment of alternative interventions to combat the spread of antimalarial resistance should be prioritised.

## Methods

### *P. falciparum* transmission model

In this study, we employed a previously developed individual-based mathematical model of *P. falciparum* malaria transmission, *magenta*, which was designed to characterise neutral genetic diversity in malaria parasites^23^ and extended to simulate the selection of antimalarial resistance.^19^ The model is an extended version of one of the models used to cost the WHO Global Technical Strategy 2030,^25^ which has been parameterized through fitting to entomological inoculation rate (EIR), parasite prevalence, clinical disease incidence, and severe disease incidence data.^24^ The model includes individual humans, mosquitoes, and parasite clones, with parasite clones represented by a genetic barcode that tracks the resistance genotype of each parasite. The extended *magenta* model has been shown to accurately capture the dynamics of complexity of infection in different malaria endemic settings,^23^ and, with two other resistance models, was used in a consensus exercise and shown to capture the speed at which resistance has been selected for.^19,26^ Full model details are provided in the appendix.

### Mapping antimalarial resistance

We tracked markers relating to resistance to artemisinin, lumefantrine, amodiaquine and piperaquine, which are the most common antimalarials taken across Africa. To map antimalarial resistance frequency for each resistance marker, we implemented a method previously used to map partner drug resistance in Africa at the first-administrative level in all malaria-endemic regions.^20^ Full details are provided in the appendix. In overview, we collated antimalarial drug resistance data in Africa from three databases: WHO Threat Maps,^27^ the WorldWide Antimalarial Resistance Network (WWARN),^28^ and the Pf7k database.^29^ These sources provided detailed information on *P. falciparum* resistance markers, including specific loci in the *pfcrt*, *pfmdr1*, *pfkelch13* genes, and copy number variations (CNV) in the *pfmdr1* and *pfpm2-3* genes. We categorised CNVs as binary variables to distinguish between single and multiple copies, giving 64 potential genotypes to be tracked, which have previously been used to infer the 28-day treatment failure for AL, ASAQ, and DHAPPQ.^30^

We deduplicated entries across databases and cleaned the resultant data to account for mixed infections and clear data entry errors. Similar to Enhrlich et al.,^20^ we split the mutation frequency data into discrete time periods and calculated the sample-weighted frequency of each resistance marker in each first-administrative region. We fit a spatial multivariate logistic regression model, incorporating spatially structured random effects through a conditional autoregressive model, to the collated resistance data to predict the frequency of drug resistance. Covariates included malaria prevalence, healthcare access, effective treatment coverage, AL pressure, seasonality, and national GDP. We used a Bayesian Markov chain Monte Carlo (MCMC) algorithm to fit the model, with 4 chains run in parallel for 10,000 burn-in iterations followed by 100,000 sampling iterations. We ensured model convergence and quality before drawing 1000 samples from the MCMC chain to construct median and 95% credible intervals for the frequency of each resistance marker. Full details are provided in the appendix.

### Modelling the spread of artemisinin resistance

To model the spread of resistance, we estimated the selection coefficient (the annual % change in logit genotype frequency^26^) for resistance frequency for the six resistance genotypes collated, assuming constant selection over time. We estimate selection coefficients using the following approach, which is described in full in the appendix.

The main drivers of antimalarial resistance in the *magenta* model are malaria transmission intensity, effective treatment coverage (the probability of an individual receiving effective treatment after developing a clinical malaria infection), the initial frequency of resistance, and the proportion of different ACTs used. We used an adjusted Latin Hypercube Sampling scheme to create 1,250 unique sets of these *magenta* model input parameters, which span the range observed globally for each parameter. For all parameter combinations, we simulated ten stochastic realisations of 100,000 individuals for 40 years to reach equilibrium first, before simulating the selection of antimalarial resistance over the following 40 years, assuming that *magenta* simulation parameters remained constant throughout. For each simulation, we calculated the selection coefficient associated with the increase in frequency of each of the six resistance genotypes (**Figure 1**).^26^

**Figure 1.**
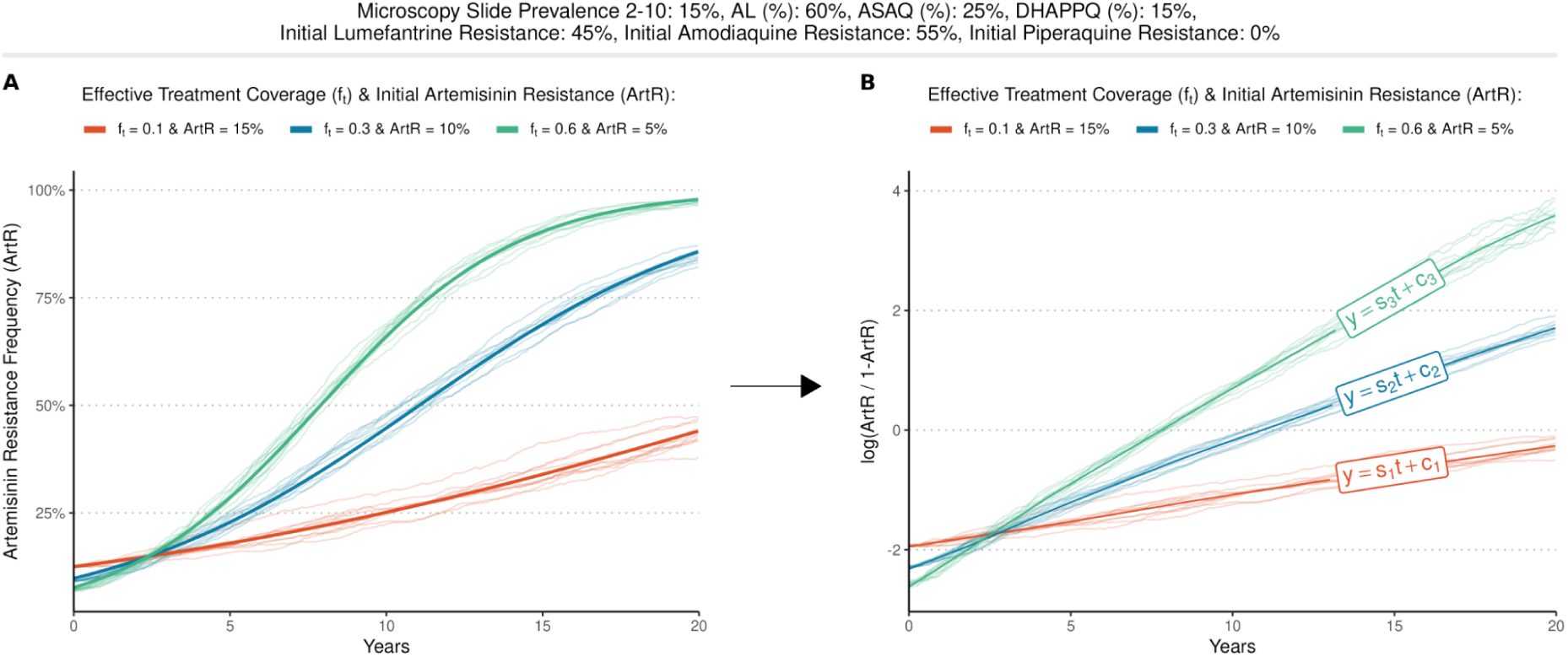
Conversion from model simulations to selection coefficients. For a given parameter set of the main drivers of resistance selection (malaria slide prevalence 2-10: 15%, percentage of front line treatment that is AL, ASAQ and DHAPPQ: 60%, 25% and 15%, initial lumefantrine, amodiaquine and piperaquine resistance: 45%, 55% and 0%), (**A**) the simulated frequency of artemisinin resistance (ART-R) is shown and is converted to (**B**) log odds (ART-R/1-ART-R), with the selection coefficient given by the calculated gradient (s_1_, s_2_, s_3_).

We trained an ensemble machine learning model to predict selection coefficients based on the *magenta* simulation parameters, yielding a statistical model that emulates the underlying transmission model behaviour and can be subsequently generalised to any malaria setting in Africa. We tested and confirmed the out-of-sample performance of the emulator, before leveraging it to estimate the selection coefficient for each resistance marker in all malaria endemic first-administrative regions in Africa. Given significant uncertainty in parameter estimates of the main drivers of antimalarial resistance, we also used the emulator to estimate an upper and lower estimate of selection coefficients, reflecting the 95% confidence interval range for each *magenta* simulation parameter. Finally, we checked the validity of the emulator to recreate the dynamics of emerging ART-R through a statistical comparison against empirical estimates of selection coefficients calculated previously in Uganda using longitudinal surveillance of ART-R.^17^

We used the estimated selection coefficients to simulate the continued spread of resistance in Africa, with the initial frequency of resistance genotypes given by the earlier mapping of resistance in Africa. To capture uncertainty in both the initial resistance frequency and the selection coefficients arising from epidemiological parameter uncertainty, we also modelled an upper and lower scenario. Initial resistance frequency was set to the 95% credible interval range from our inferred resistance maps, and selection coefficients were estimated based on the 95% confidence interval range for *magenta* simulation parameters.

Given the difficulty in estimating the rate at which malaria parasites under selection spread geographically,^31^ we used a simple model of parasite movement in space. Resistant parasites spread out from a source region to all adjacent first-administrative regions at an equal rate, with the rate determined such that once resistance in the source region reaches 25% frequency, resistance in all neighbouring regions is established and not susceptible to stochastic fade-out. Once resistance is established in a neighbouring region (defined as 1% resistance frequency based on previous antimalarial resistance consensus modelling exercises <u>(Watson et al. 2022)</u>), we assume all future resistance dynamics are solely determined by the region’s selection coefficient. Given the use of a single fixed selection coefficient for each region, this assumes that malaria prevalence and case management in each region remains constant over time. Using this approach, the resultant range in possible timelines for the spread of antimalarial resistance in Africa is based only on uncertainty in current resistance frequency and epidemiological parameters, with a single model of spatial spread and no assumed changes in future case management or malaria control. We used the projected spread of resistance mutations in Africa to infer the increase in the probability of 28-day treatment failure in each first-administrative region, before using the projected annual population size in each region to estimate population-weighted treatment failures in Africa till 2060.^32^

## Results

After cleaning, deduplication, and accounting for mixed infections, we identified 3,808 surveys of resistance collected from 2000 to 2021 in Africa, with a mean number of samples tested of 65. Surveys provided estimates of the resistance frequency for the K76T locus in *pfcrt*, the N86Y and Y184F loci in *pfmdr1*, WHO validated markers of artemisinin resistance in *pfkelch13* and copy-number variation (CNV) of *pfmdr1*, and CNV of *pfpm2-3* (Supplementary Figure 1). There was considerable spatial and temporal sparsity in the availability of molecular surveys, with a notable decline in the availability of molecular surveys in recent years, likely reflecting the delay from sample collection to molecular surveillance data being published. Analysis of the spatial distribution of all resistance markers (Supplementary Figures 2-7) revealed significant spatial autocorrelation (p < 0.05) for all markers except *pfmdr1* N86Y. Across Africa, the mean frequency of resistance was highest for *pfcrt* K76T and lowest for combined *pfkelch13* WHO validated markers of ART-R.

From the geostatistical models fit to each marker, we found a significant predictive effect for all covariates. Specifically focusing on ART-R, we found a positive correlation between ART-R frequency and both effective treatment coverage and the proportion of ACTs administered that were AL. Conversely, factors such as higher malaria prevalence, better healthcare access, and higher GDP were associated with reduced ART-R. The resultant inferred map of ART-R frequency in Africa confirmed the high spatial autocorrelation, with the 50% credible interval map indicating limited geographic spread of ART-R, with notable exceptions seen in the known current hotspots of emerging ART-R in Uganda, Rwanda and Eritrea (Figure 2). The inferred maps of resistance frequency for ART-R and the other fiver markers of resistance (Supplementary Figures 8-13) were subsequently used to characterise the risk of resistance selection by estimating selection coefficients for each marker using the emulator trained on simulated model outputs for the trajectory of resistance (Supplementary Figure 14), before conducting forward simulations to project future trends in antimalarial resistance and treatment failure.

**Figure 2.**
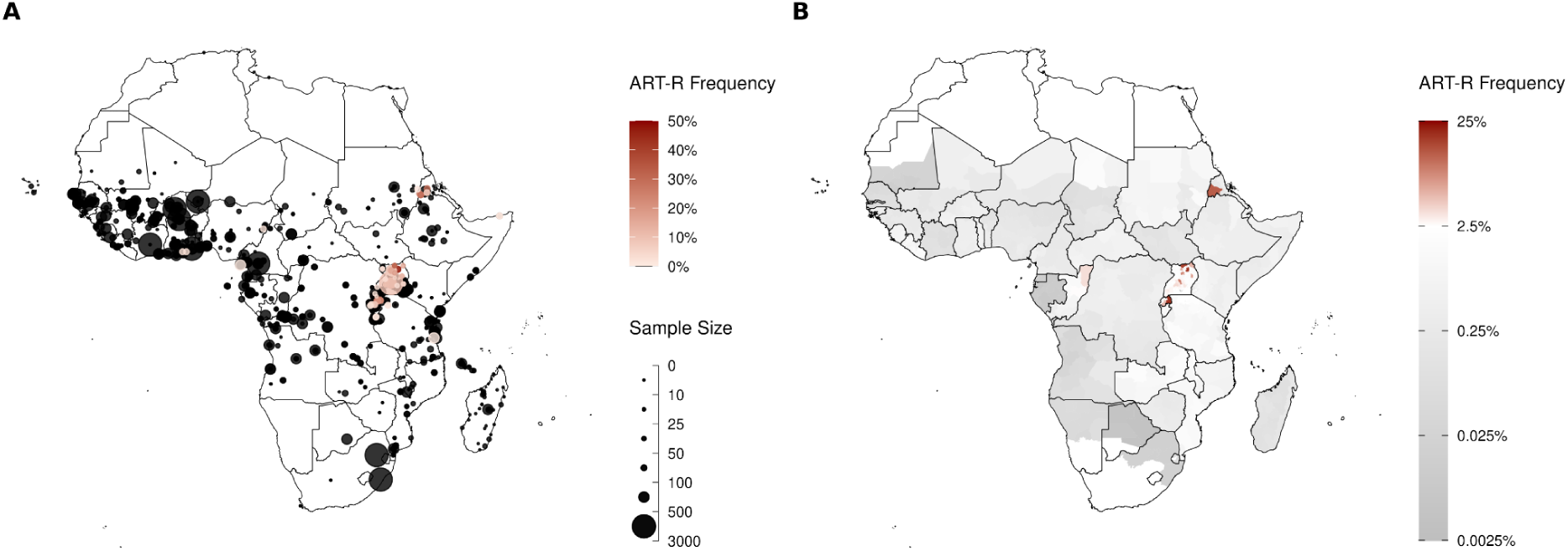
Frequency of WHO validated *pfkelch13* markers of artemisinin resistance. **(A)** Each point shows an individual molecular survey for *pfkelch13* mutations collected between 2017-2021 in Africa. Point size shows the number of samples tested, and increasing frequency of WHO validated *pfkelch13* markers of artemisinin resistance (ART-R) is shown in shades of red. Any survey in which no ART-R was observed is shown in black. **(B)** Map shows the 50% credible interval estimate from a Bayesian spatial model of ART-R frequency at the first-administrative region. Any region predicted to have greater than 2.5% ART-R frequency is shown in shades of red. See **appendix** for equivalent maps and 95% credible intervals for all other genetic markers modelled.

A single selection coefficient over time was able to capture the dynamics of resistance in the transmission model output for all the six genotypes of interest. Using our trained emulator, we mapped the distribution of selection coefficients for ART-R, revealing significant variation across Africa (Figure 3). For the selection of ART-R, across all first-administrative regions, our central estimates of selection coefficients ranged from 0.059 - 0.404, representing 5.9% - 40% relative increase in resistance frequency per year (Figure 32A). We observed the highest selection coefficients, indicating a rapid increase in resistance frequency, in East Africa, particularly in regions surrounding the current epicentre of emerging ART-R in Rwanda and Uganda. We used these selection coefficients to estimate the corresponding time for the ART-R to increase from 1% to 10% frequency, which ranged between 5.9 - 40.7 years (Figure 3B). An analysis of the partial predictive plots for the emulator showed that the spatial pattern observed in the mapped selection coefficients was driven predominantly by high effective treatment coverage, with low malaria prevalence and low levels of multiple ACTs being used, as determined by a higher proportion of all ACTs administered being AL, driving higher selection coefficients (Supplementary Figure 15).

**Figure 3.**
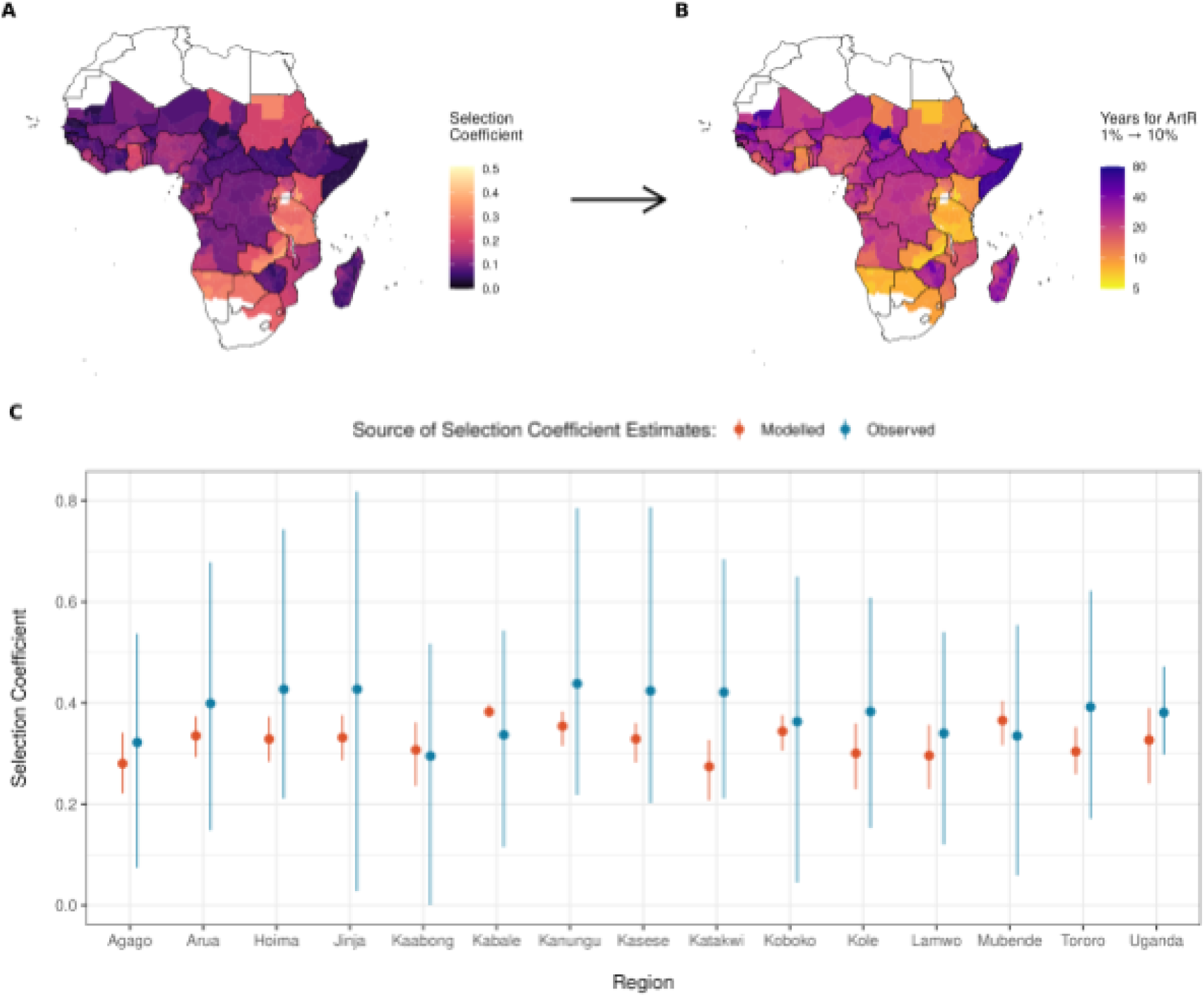
Risk of artemisinin selection in Africa. **A)** Mapped distribution of selection coefficients for artemisinin resistance in Africa and **B)** the resultant modelled time in years for artemisinin resistance (ART-R) to increase from 1% to 10% frequency. **C)** Out of sample comparison of model predicted selection coefficients (red) for Uganda against directly estimated selection coefficients (blue) from Meier-Scherling et al. (2024). Points and whiskers in blue represent median and 95% CrI from Meier-Scherling et al. (2024) and points and whiskers in red show the central, upper and lower estimates based on the median and 95% CI estimates for model simulation parameters.

We tested the validity of the developed emulator model by comparing the predicted selection coefficients in Uganda against those previously estimated directly using longitudinal molecular surveillance data between 2016-2022. The model predictions for Uganda closely matched the empirical estimates, with the modelled central estimate for each region included in the 95% confidence interval estimated from longitudinal surveillance data for each region and nationally (Figure 3C). Overall, the model-predicted selection coefficients were lower than those estimated directly from the observed data. However, they were a significantly better fit to the observed data than the predicted selection coefficients estimated when non-malarial fevers were not accounted for in the model (Supplementary Figure 16). When accounting for uncertainty in parameter estimates of the main drivers of antimalarial resistance, the mean increase and decrease in selection coefficients across all regions was 13.9% (95% CI: 4.3% - 30.9%) and 18.4% (95% CI: 4.7% - 44.0%) for the upper and lower estimates respectively (Supplementary Figure 17).

If current malaria prevalence and treatment coverage and drug policies remain constant, our scenario projections predict that ART-R will continue to spread across Africa over the next 20 years (Figure 4). The model predicts that resistance will continue to spread from the current hotspots in Rwanda and Uganda, with significant expansion into neighbouring countries such as Kenya and Tanzania. In our central scenario, we predict that ART-R will be greater than 10% frequency by 2040 in the majority of first-administrative regions in 16 countries, mostly concentrated in East Africa. When accounting for uncertainty in parameter estimates of the main drivers of antimalarial resistance, we predict that ART-R will be greater than 10% frequency by 2040 in 11 and 26 countries based on the lower and upper parameter estimate uncertainty (Supplementary Figure 18).

**Figure 4.**
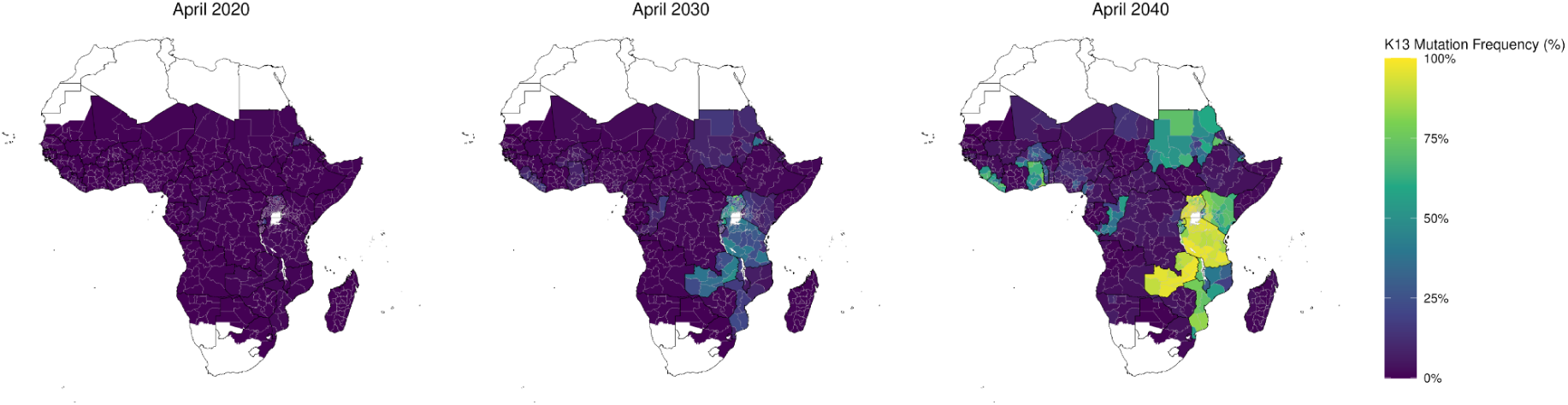
Spread of artemisinin resistance frequency in Africa. The central estimate for the predicted spread of artemisinin resistance frequency (ART-R) in Africa during 2020 - 2040. ART-R is predicted to spread out from the current epicentres of artemisinin resistance in Rwanda and Uganda, with 16 countries predicted to have greater than 10% ART-R by 2040 in the majority of first-administrative regions.

In our modelled scenarios of the spread of ART-R, the fastest selection of ART-R occurs in settings that also select quickly for the *pfmdr1* Y184F mutation and *pfmdr1* CNV, which confers an increased tolerance to lumefantrine. The resultant selection of both ART-R and decreasing lumefantrine efficacy leads to decreases in the average efficacy of AL and increases in the percentage of 28-day treatment failures across Africa. Using our central estimates for the speed of selection, we predict that the mean percentage of treatment failures across Africa, weighted by predicted population sizes in 2060 and based on current clinical incidence rates per capita, will reach 30.74% by 2060 (Figure 5, Table 1). When accounting for uncertainty in parameter estimates of the main drivers of antimalarial resistance, our estimates range between 24.98% - 34.54% for the lower and upper parameter estimate uncertainty, respectively. Using current estimates of effective-treatment coverage, this translates to 52,980,600 absolute occurrences of treatment failure in 2060, ranging between 26,374,200 - 93,672,400 for the lower and upper parameter estimate uncertainty, respectively.

**Figure 5.**
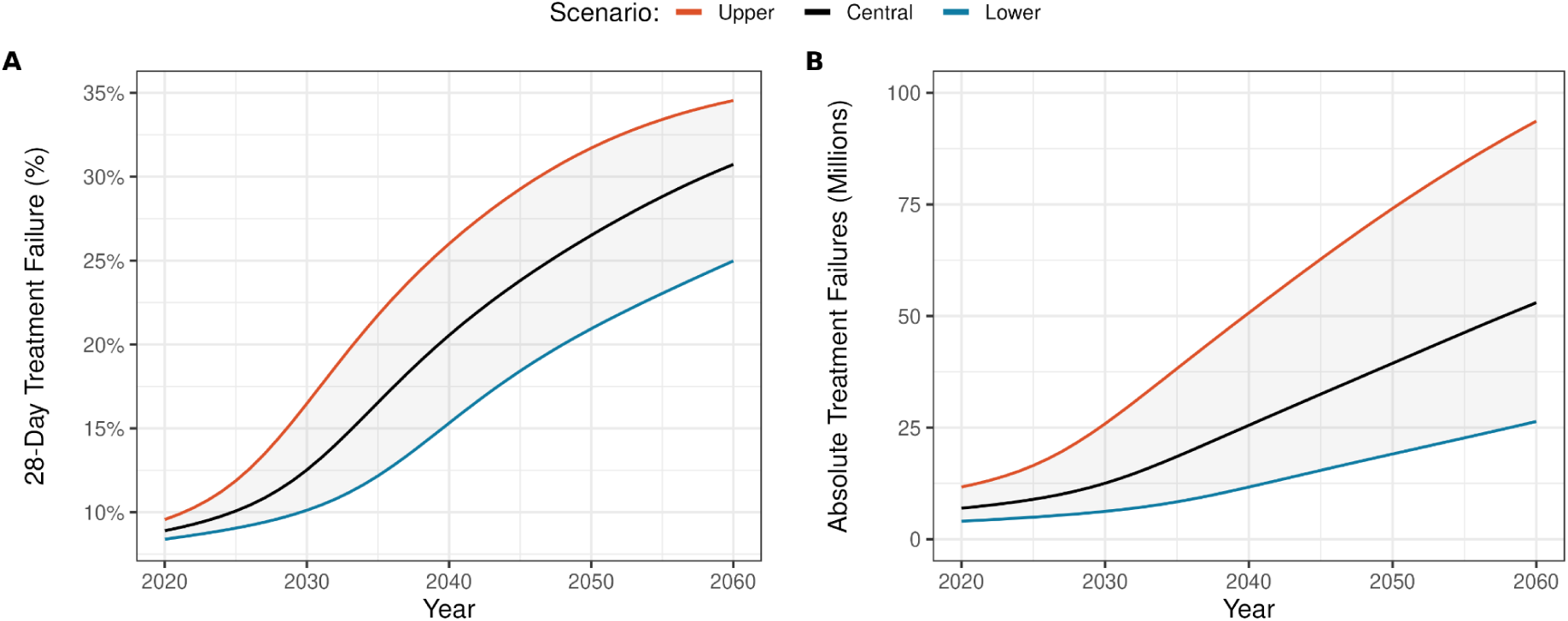
Projected treatment failure in Africa without changes in drug policy. **(A)** The modelled increase in treatment failure and **(B)** absolute number of treatment failures over the next 40 years under the upper, central and lower scenarios, where upper and lower scenarios reflect the 95% CI uncertainty in parameter estimates of the main drivers of antimalarial resistance.

**Table 1.**
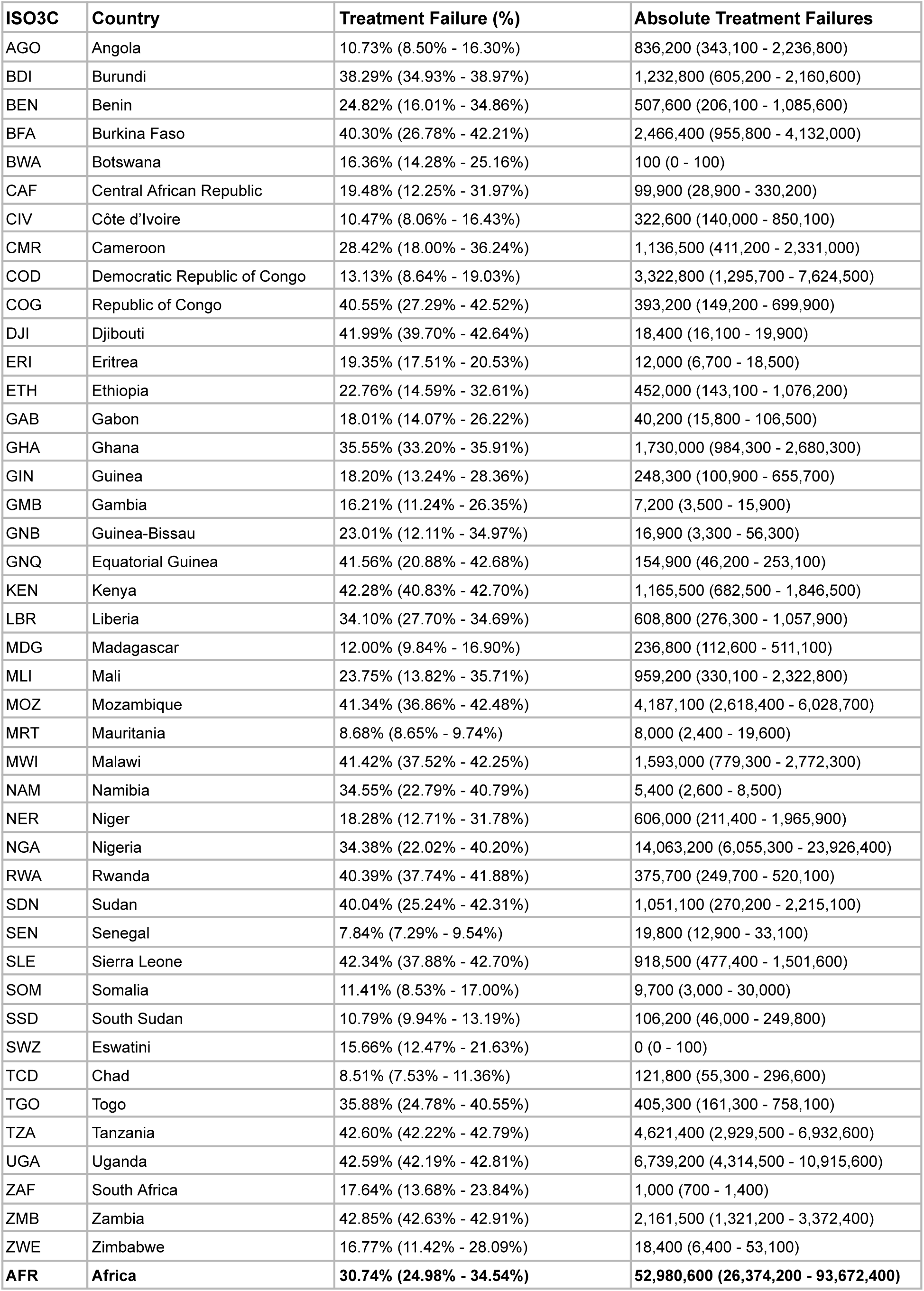
Average Treatment Failure and Absolute Number of Treatment Failures in Africa in 2060.

| **ISO3C** | **Country** | **Treatment Failure (%)** | **Absolute Treatment Failures** |
| --- | --- | --- | --- |
| AGO | Angola | 10.73% (8.50% - 16.30%) | 836,200 (343,100 - 2,236,800) |
| BDI | Burundi | 38.29% (34.93% - 38.97%) | 1,232,800 (605,200 - 2,160,600) |
| BEN | Benin | 24.82% (16.01% - 34.86%) | 507,600 (206,100 - 1,085,600) |
| BFA | Burkina Faso | 40.30% (26.78% - 42.21%) | 2,466,400 (955,800 - 4,132,000) |
| BWA | Botswana | 16.36% (14.28% - 25.16%) | 100 (0 - 100) |
| CAF | Central African Republic | 19.48% (12.25% - 31.97%) | 99,900 (28,900 - 330,200) |
| CIV | Côte d'Ivoire | 10.47% (8.06% - 16.43%) | 322,600 (140,000 - 850,100) |
| CMR | Cameroon | 28.42% (18.00% - 36.24%) | 1,136,500 (411,200 - 2,331,000) |
| COD | Democratic Republic of Congo | 13.13% (8.64% - 19.03%) | 3,322,800 (1,295,700 - 7,624,500) |
| COG | Republic of Congo | 40.55% (27.29% - 42.52%) | 393,200 (149,200 - 699,900) |
| DJI | Djibouti | 41.99% (39.70% - 42.64%) | 18,400 (16,100 - 19,900) |
| ERI | Eritrea | 19.35% (17.51% - 20.53%) | 12,000 (6,700 - 18,500) |
| ETH | Ethiopia | 22.76% (14.59% - 32.61%) | 452,000 (143,100 - 1,076,200) |
| GAB | Gabon | 18.01% (14.07% - 26.22%) | 40,200 (15,800 - 106,500) |
| GHA | Ghana | 35.55% (33.20% - 35.91%) | 1,730,000 (984,300 - 2,680,300) |
| GIN | Guinea | 18.20% (13.24% - 28.36%) | 248,300 (100,900 - 655,700) |
| GMB | Gambia | 16.21% (11.24% - 26.35%) | 7,200 (3,500 - 15,900) |
| GNB | Guinea-Bissau | 23.01% (12.11% - 34.97%) | 16,900 (3,300 - 56,300) |
| GNQ | Equatorial Guinea | 41.56% (20.88% - 42.68%) | 154,900 (46,200 - 253,100) |
| KEN | Kenya | 42.28% (40.83% - 42.70%) | 1,165,500 (682,500 - 1,846,500) |
| LBR | Liberia | 34.10% (27.70% - 34.69%) | 608,800 (276,300 - 1,057,900) |
| MDG | Madagascar | 12.00% (9.84% - 16.90%) | 236,800 (112,600 - 511,100) |
| MLI | Mali | 23.75% (13.82% - 35.71%) | 959,200 (330,100 - 2,322,800) |
| MOZ | Mozambique | 41.34% (36.86% - 42.48%) | 4,187,100 (2,618,400 - 6,028,700) |
| MRT | Mauritania | 8.68% (8.65% - 9.74%) | 8,000 (2,400 - 19,600) |
| MWI | Malawi | 41.42% (37.52% - 42.25%) | 1,593,000 (779,300 - 2,772,300) |
| NAM | Namibia | 34.55% (22.79% - 40.79%) | 5,400 (2,600 - 8,500) |
| NER | Niger | 18.28% (12.71% - 31.78%) | 606,000 (211,400 - 1,965,900) |
| NGA | Nigeria | 34.38% (22.02% - 40.20%) | 14,063,200 (6,055,300 - 23,926,400) |
| RWA | Rwanda | 40.39% (37.74% - 41.88%) | 375,700 (249,700 - 520,100) |
| SDN | Sudan | 40.04% (25.24% - 42.31%) | 1,051,100 (270,200 - 2,215,100) |
| SEN | Senegal | 7.84% (7.29% - 9.54%) | 19,800 (12,900 - 33,100) |
| SLE | Sierra Leone | 42.34% (37.88% - 42.70%) | 918,500 (477,400 - 1,501,600) |
| SOM | Somalia | 11.41% (8.53% - 17.00%) | 9,700 (3,000 - 30,000) |
| SSD | South Sudan | 10.79% (9.94% - 13.19%) | 106,200 (46,000 - 249,800) |
| SWZ | Eswatini | 15.66% (12.47% - 21.63%) | 0 (0 - 100) |
| TCD | Chad | 8.51% (7.53% - 11.36%) | 121,800 (55,300 - 296,600) |
| TGO | Togo | 35.88% (24.78% - 40.55%) | 405,300 (161,300 - 758,100) |
| TZA | Tanzania | 42.60% (42.22% - 42.79%) | 4,621,400 (2,929,500 - 6,932,600) |
| UGA | Uganda | 42.59% (42.19% - 42.81%) | 6,739,200 (4,314,500 - 10,915,600) |
| ZAF | South Africa | 17.64% (13.68% - 23.84%) | 1,000 (700 - 1,400) |
| ZMB | Zambia | 42.85% (42.63% - 42.91%) | 2,161,500 (1,321,200 - 3,372,400) |
| ZWE | Zimbabwe | 16.77% (11.42% - 28.09%) | 18,400 (6,400 - 53,100) |
| **AFR** | **Africa** | **30.74% (24.98% - 34.54%)** | **52,980,600 (26,374,200 - 93,672,400)** |

## Discussion

In this study, we modelled the risk of selection and spread of ART-R in Africa, providing a detailed assessment of how resistance could evolve and expand over the coming decades. Our findings indicate that the highest selection coefficients for ART-R are predominantly located in regions with high effective treatment coverage with predominantly one ACT, low malaria prevalence and high partner drug resistance. In particular, our model framework correctly identified the current epicentres of ART-R in Rwanda and Uganda and surrounding regions as those with the highest selection coefficient for ART-R, with the selection coefficients inferred in Uganda consistent with those measured directly from longitudinal molecular surveillance.^17^ In forward scenario projections without changes in drug policy, we predict that ART-R will exceed 10% frequency by 2040 in sixteen African countries, concentrated in East Africa near the current epicentre of ART-R in Rwanda and Uganda. The resultant reduction in treatment efficacy could lead to greater than 30% 28-day treatment failure rates on average in Africa by 2060, resulting in tens of millions of treatment failures by 2060.

The implications of these findings for malaria control are profound. The projected rise in treatment failures due to ART-R highlighted in our projections signals a growing public health crisis. If current treatment policies remain unchanged, treatment failure rates could escalate above the WHO treatment failure threshold of 10%, severely undermining our ability to effectively treat malaria cases, reducing and leading to a resurgence in malaria morbidity and mortality. The current reliance on artemisinin in combination therapies and clear potential for ART-R to spread requires immediate containment strategies in high-risk areas. Firstly, there is an urgent need to strengthen molecular surveillance systems across Africa to detect and monitor the spread of ART-R.^5^ Current surveillance efforts are concentrated in a few regions, with our spatial mapping of antimalarial resistance markers showing large regions of the continent under-monitored, significantly limiting the precision in our efforts to map antimalarial resistance. Expanding molecular surveillance to regions without recent and comprehensive surveillance, particularly those we identified with high selection coefficients for ART-R, is essential to improve mapping efforts and providein providing sufficiently early detection of ART-R for effective containment strategies to be implemented. Molecular surveillance must also include partner drug resistance markers and be integrated with routine malaria case management monitoring.^33^ We also need additional and more flexible early warning tools to detect increasing ACT tolerance. Monitoring the proportion of treated malaria patients returning to clinics with recurrent infections,^11^ the time to reinfection after treatment^34^ and the therapeutic efficacy in imported malaria cases in non-endemic regions^12^ all provide additional lenses to detect decreasing ACT susceptibility earlier.

The close agreement between our modelled selection coefficients and those directly measured in Uganda^17^ provides increased confidence that our modelling framework reflects the current emergence of ART-R in Uganda. However, our central estimates of the speed of selection of ART-R in Ethiopia are very low, due to low effective-treatment coverage estimates, and likely underestimate the risk of ART-R selection given the emergence of 662I mutations. These contrasting findings, however, could be explained by the considerable uncertainty in our modelled scenarios and why the full range of uncertainty presented should be considered. Much of the uncertainty is due to uncertainty in our subnational estimates of malaria prevalence, effective treatment coverage, the relative use of different ACTs and current levels of antimalarial resistance. This uncertainty causes our upper estimate of the absolute number of treatment failures (93,672,400) to be over three and half times larger than our lower estimate (26,374,200). Improving estimates and our understanding of each of these factors is necessary to improve the accuracy and precision of modelling, while also enabling the design of region-specific public health policies to combat resistance.

In an effort to provide a comprehensive assessment of resistance across Africa, we introduced various limiting assumptions in our study. While we characterised the impact of uncertainty in a number of key drivers of resistance, there are other drivers that were not explored as extensively in our sensitivity analyses. Our results show that accounting for individuals with asymptomatic malaria infection seeking treatment for a non-malarial febrile illness increases ART-R selection. However, current estimates of subnational and age-disaggregated estimates of the frequency of these events are limited.^36^ The impact of the private drug market towards resistance is poorly understood. While our estimates of effective treatment coverage do account for the private market, the quality of private market drugs may be lower than in the public sector. Similarly, incomplete drug adherence is also not explicitly modelled, although substantial evidence points towards significant heterogeneity in adherence.^13^ Furthermore, while the spatial model of parasite movement to simulate the geographic spread of resistance is a practical simplification, it does not fully account for the complex human and environmental factors that influence parasite migration and gene flow across regions. Lastly, the assumption of constant malaria transmission intensity and treatment coverage over time is clearly an incorrect assumption. However, it is helpful nonetheless in providing a counterfactual for the possible spread of resistance if changes in treatment policy, such as MFT or novel non-artemisinin combination therapies such as ganaplacide-lumefantrine, are not implemented.

Despite these limitations, the generated results, including spatial estimates of current resistance frequency, identification of regions most likely to select for resistance once established and scenario timelines for further resistance spread under current drug policies, will help guide future efforts to combat antimalarial resistance. Further, it is clear that ART-R resistance is emerging and increasing in Africa and policy makers will need to consider alternative strategies to combat resistance. Widespread availability of ganaplacide-lumefantrine is likely still five years away with Phase 3 trials ongoing after successful Phase 2 results,^37^ which will reduce selective pressures on artemisinin but is reliant on lumefantrine remaining fully effective. Triple artemisinin combination therapies are a particularly promising tool to combat resistance, reinforcing the effectiveness of combining drugs in a single formulation and removing the time window between parasites being exposed to different drugs.^38^ Before these new tools are widespread, strategic use of multiple first-line therapies will likely be needed to limit the selection of resistance, which will require careful consideration of the logistical challenges of distributing multiple treatments across diverse and resource-constrained settings. Importantly, each of these changes in treatment policies will not prevent resistance unless resistance drivers, such as incomplete drug adherence, lack of diagnostic testing and private marker drug quality are addressed. As with many areas of malaria control, it is likely that multiple tools, evolving over time to keep pace with parasite evolution, will be needed to prevent resistance, reversing the last two decades of reductions in malaria burden.

### Contributions

OJW conceived the study with input from SM, AW, GS and AMW. OJW led the modelling and statistical analysis, with input from TB, GC-D, PW and LO. CM-S, VA, TK and JAB contributed data and interpretation of model comparisons to data in Uganda. OJW produced the first draft of the manuscript and has accessed and verified the underlying data. All authors read, contributed to, and approved the final draft. All authors had full access to all the data in the study and had final responsibility for the decision to submit for publication.

### Data sharing

All data, codes, and supplementary tables used and generated by this study are available in a <u>Github Repository</u> (version 1.0.1). All estimates of resistance spread are in the GitHub Repository.

## Data Availability

All data, codes, and supplementary tables used and generated by this study are available in a Github Repository (version 1.0.1). All estimates of resistance spread are in the GitHub Repository.

## Acknowledgements

OJW is supported by an Imperial College Research Fellowship sponsored by Schmidt Sciences. OJW, LO, CM-S and JAB were supported by US NIH (R01AI156267). PW acknowledges support from the Bill & Melinda Gates Foundation (INV-043624).

## Supplementary Material

Supplementary Appendix Attached

