## Supplementary Appendix for "Risk of selection and timelines for the continued spread of artemisinin and partner drug resistance in Africa"

—-----------------------------------------------------------------------------------------------------------

*For the purpose of open access, the author has applied a ‘Creative Commons Attribution (CC BY) licence (where permitted by UKRI, ‘Open Government Licence’ or ‘Creative Commons Attribution No-derivatives (CC-BY-ND) licence’ may be stated instead) to any Author Accepted Manuscript version arising.*

—-----------------------------------------------------------------------------------------------------------

### Supplementary Methods

#### *P. falciparum* transmission model

We used a previously developed individual-based stochastic model to simulate the transmission dynamics of *Plasmodium falciparum*. The model was first developed to model the patterns in neutral genetic diversity (Watson et al. 2021) and was extended to model the selection of antimalarial resistance.[1] Briefly, it is an individual based, discrete time, stochastic model, with individual mosquitoes explicitly modelled, originally developed for characterising the impact of transmission intensity on neutral genetic diversity.

Both the human and adult mosquito stages are modelled at an individual level, whereas parasites are modelled as discrete populations with each population relating to an infection event. The human transmission model is based upon previous modelling efforts,[2–4] which is described in its deterministic framework first, before detailing the human acquisition of immunity and the full set of equations detailing its stochastic implementation. The deterministic model described within the methods has been included as its equilibrium solution is used for model initialisation. Additionally, we developed a deterministic version of the earlier 2016 Griffin et al. model[4] that incorporates interventions, which is used to indirectly incorporate the effects of intervention strategies as these are not modelled explicitly within the individual model. (The deterministic implementation of interventions has not been included within the deterministic model described below to ensure clarity related to our indirect handling of interventions).

We continue to describe the mosquito transmission model, which is again based on earlier modelling efforts,[2–4] before describing the stochastic equations detailing the implementation of the adult mosquito stage at an individual-based level. We follow by describing how the parasite populations are modelled by using a genetic barcode to encode the resistance genotype of each parasite.[1] We continue by describing the within-host parasite populations, which includes considerations surrounding the contribution of coinfection and superinfection towards the model's dynamics of within-host multiplicities of infection, and how these relate to the probabilistic uptake of specific gametocyte strains by mosquitoes.[5] This is followed by detailing the within-mosquito parasite populations, which explores the derivation of the distribution describing the model-predicted oocyte intensities, and describes how recombination within the sexual stage is explicitly modelled.

##### Human Transmission Model

###### Human Transmission

Individuals begin life susceptible to infection (state S) (**Supplementary Figure 1**). At birth, individuals possess a level of maternal immunity that decays exponentially over the first 6 months. Each day individual $i$ is probabilistically exposed to infectious bites governed by their individual force of infection ($\Lambda_i$). $\Lambda_i$ is dependent on their pre-erythrocytic immunity, exposure to bites (dependent on both their age and their individual relative biting rate due to heterogeneous biting patterns in mosquitoes) and the size of the infectious mosquito population. Infected individuals, after a latent period of 12 days ($d_E$), develop either clinical disease (state D) or asymptomatic infection (state A). This outcome is determined by their probability of acquiring clinical disease ($\phi_i$), which is dependent on their clinical immunity. Individuals who develop the disease have a fixed probability ($f_T$) of seeking treatment (state T). Treated individuals are assumed to always recover, i.e. fully-curative treatment, and then enter a protective state of prophylaxis (state P) at rate $r_T$, before returning to susceptible at rate $r_s$. Individuals who did not receive treatment recover to a state of asymptomatic infection at rate $r_D$. Asymptomatic individuals progress to a subpatent infection (stage U) at rate $r_A$, before clearing infection and returning to susceptible at rate $r_U$. Additionally, superinfection is possible for all individuals in states A and U. Superinfected individuals who receive treatment will move to state T. Individuals who are superinfected but do not receive treatment in response to the superinfection will either develop clinical disease, thus moving to state D, or develop an asymptomatic infection and move to state A.

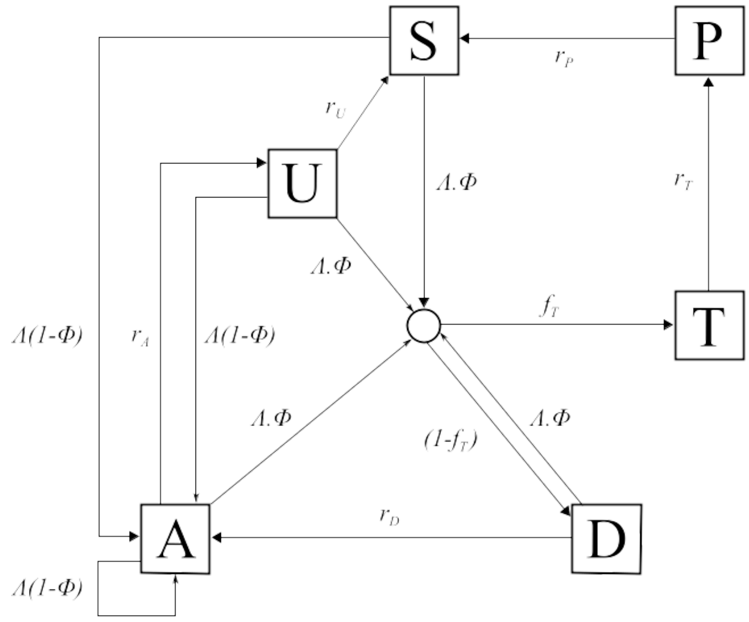


**Supplementary Methods Figure 1: Transmission Model.** Flow diagram for the human component of the transmission model, with dashed arrows indicating superinfection. S, susceptible; T, treated clinical disease; D, untreated clinical disease; P, prophylaxis; A, asymptomatic patent infection; U, asymptomatic sub-patent infection. All parameters are described and referenced within Supplementary Methods Table 1.

The movement between the human components of the transmission model is summarised with the following partial differential equations describing each compartment (*t* represents time and *a* represents age):

$$\frac{\partial S}{\partial t} + \frac{\partial S}{\partial a} = -\Lambda(t - d_E)S + \frac{P(t)}{d_P} + \frac{U(t)}{d_U}$$

$$\frac{\partial T}{\partial t} + \frac{\partial T}{\partial a} = \phi f_T \Lambda(t - d_E)(S(t) + A(t) + U(t)) - \frac{T(t)}{d_T}$$

$$\frac{\partial D}{\partial t} + \frac{\partial D}{\partial a} = \phi(1 - f_T)\Lambda(t - d_E)(S(t) + A(t) + U(t)) - \frac{D(t)}{d_D}$$

$$\frac{\partial A}{\partial t} + \frac{\partial A}{\partial a} = (1 - \phi)\Lambda(t - d_E)(S(t) + U(t)) + \frac{D(t)}{d_D} - \phi\Lambda A(t) - \frac{A(t)}{d_A}$$

$$\frac{\partial U}{\partial t} + \frac{\partial U}{\partial a} = \frac{A(t)}{d_A} - \frac{U(t)}{d_U} - \Lambda(t - d_E)U(t)$$

$$\frac{\partial P}{\partial t} + \frac{\partial P}{\partial a} = \frac{T(t)}{d_T} - \frac{P(t)}{d_P}$$

When an individual enters a new infection state a waiting time is sampled from an exponential distribution for when the individual will move out of that infection state (except when individuals move into S). With the introduction of a fixed daily time-step, the day on which an individual transitions from state X to Y occurs is given by:

$$Day(X \rightarrow Y) \sim floor(Exp(\lambda)) + t_{now} + 1$$

where $t_{now}$ is the current day, i.e. the day that the individual moved into state A, and λ is the transition rate. The set of state transitions for individuals and their associated transition rates are given below.

| Process | Transition | Transition Rate |
|---|---|---|
| Progression of untreated disease to asymptomatic infection | D → A | $r_D = \frac{1}{d_D}$ |
| Progression of asymptomatic infection to subpatent infection | A → U | $r_A = \frac{1}{d_A}$ |
| Progression of subpatent infection to susceptible | U → S | $r_U = \frac{1}{d_U}$ |
| Progression of treated disease to uninfected prophylactic period | T → P | $r_T = \frac{1}{d_T}$ |
| Progression from uninfected prophylactic period to susceptible | P → S | $r_P = \frac{1}{d_P}$ |

We assume that each person has a unique biting rate, which is the product of their relative age-dependent biting rate, $\psi_i$, given by

$$\psi_i(a) = \frac{\sum_{i=1}^{n} \psi_i(a)}{n}\left(1 - \rho exp^{-\frac{a}{a_0}}\right)$$

and an assumed heterogeneity in biting patterns of mosquitoes, $\zeta_i$, which we assume persists throughout their lifetime and is drawn from a log-normal distribution with a mean of 1,

$$log\left(\zeta_i\right) \sim N\left(\frac{-\sigma^2}{2}, \sigma^2\right)$$

where $1 - \rho$ is the relative biting rate at birth when compared to adults and $a_0$ represents the time-scale at which the biting rate increases with age. The product of these biting rates is subsequently used to calculate the proportion of the whole population's bites that person *i* receives on a given day, $\pi_i$. Their daily entomological inoculation rate (EIR), $\epsilon_i$, is thus calculated by multiplying by the number of infectious mosquitoes taking a blood meal from a human that day, which in turn yields their force of infection, which are given by

$$\pi_i = \zeta_i \psi_i$$

$$\epsilon_i = I_{M\_Feeding} \pi_i$$

$$\Lambda_i = \epsilon_i b_i$$

where $I_{M\_Feeding}$ is the size of the feeding infectious mosquito population, and $b_i$ is the probability of infection given an infectious mosquito bite.

The inclusion of individual mosquitoes results in the following stochastic implementation of infection. On any given day, the number of infectious mosquitoes taking a blood meal from a human ($I_{M\_Feeding}$) will result in the same number of infectious bites. These bites are allocated by sampling from the multinomial distribution using the conditional binomial method,[6] where sample weights are equal to $\pi_i$. Upon receiving an infectious bite, an individual will move to an untracked infection state, *I,* which leads to either clinical disease (*D*), treated clinical disease (*T*), or asymptomatic infection (*A*). This leads to the following transition rates related to infection below.

| Process | Transition | Transition Rate |
|---|---|---|
| Infection | S → I | $\Lambda_i(t - d_E)$ |
| Super-infection from untreated clinical disease, asymptomatic infection or subpatent infection | D → I<br>A → I<br>U → I | $\Lambda_i(t - d_E)$ |

The probabilities of progressing from state I to D, T or U are determined by drawing a sequence of Bernoulli trials for each infected individual as:

$$Prob(Clinical\ Disease):\ Bernoulli(\phi_i)$$

$$Prob(Treated\ Clinical\ Disease\ |\ Clinical\ Disease):\ Bernoulli(f_T)$$

The human population was assumed to have a maximum possible age of 100 years, with an average age of 21 years within the population, yielding an approximately exponential age distribution typical of sub-Saharan countries. The day on which a human dies is thus allocated at birth by sampling from an exponential distribution with a mean equal to 21 years. When an individual dies, they are replaced with a new-born individual with the same individual biting rate due to heterogeneity in biting patterns.

**Immunity and Detection Functions**

We model 3 stages at which immunity may impact transmission, as in the existing Griffin et al model:

1. Pre-erythrocytic immunity, $I_B$; reduction in the probability of infection given an infectious mosquito bite.

2. Acquired and Maternal Clinical Immunity, $I_{CA}$ and $I_{CM}$ respectively; reduction in the probability of clinical disease given an infection due to the effects of blood stage immunity.

3. Detection immunity, $I_D$; reduction in the probability of detection.

Maternal clinical immunity is assumed to be at birth a proportion, $P_M$, of the acquired immunity of a 20 year-old and to decay at rate $\frac{1}{d_M}$. The remaining three types of immunity are described by the following partial differential equations, which describe how immunity increases due to exposure from zero at birth and decreases over time:

$$\frac{\partial I_B}{\partial t} + \frac{\partial I_B}{\partial a} = \frac{\epsilon}{\epsilon u_B + 1} - \frac{I_B}{d_B}$$

$$\frac{\partial I_{CA}}{\partial t} + \frac{\partial I_{CA}}{\partial a} = \frac{\Lambda}{\Lambda u_C + 1} - \frac{I_{CA}}{d_{CA}}$$

$$\frac{\partial I_D}{\partial t} + \frac{\partial I_D}{\partial a} = \frac{\Lambda}{\Lambda u_D + 1} - \frac{I_D}{d_{ID}}$$

where each $u$ term represents the time during which immunity cannot be boosted further after a previous boost and each $d$ term represents the duration of immunity.

The probabilities of infection, detection and clinical disease are subsequently created by transforming each immunity function by Hill functions. An individual's probability of infection, $b_i$, is given by

$$b_i = b_0\left(b_1 + \frac{1-b_1}{1+\left(\frac{I_B}{I_{B0}}\right)^{\kappa_B}}\right)$$

where $b_0$ is the maximum probability due to no immunity, $b_0 b_1$ is the minimum probability and $I_{B0}$ and $\kappa_B$ are scale and shape parameters respectively.

An individual's probability of clinical disease, $\phi_i$, is given by

$$\phi_i = \phi_0\left(\phi_1 + \frac{1-\phi_1}{1+\left(\frac{I_{CA}+I_{CM}}{I_{C0}}\right)^{\kappa_C}}\right)$$

where $\phi_0$ is the maximum probability due to no immunity, $\phi_1\phi_0$ is the minimum probability and $I_{C0}$ and $\kappa_C$ are scale and shape parameters respectively.

An individual's probability of being detected by microscopy when asymptomatic, $q_i$, is given by

$$q_i = d_1 + \left(\frac{1-d_1}{1+\left(\frac{I_D}{I_{D0}}\right)^{\kappa_D} f_D}\right)$$

where $d_1$ is the minimum probability due to maximum immunity, and $I_{D0}$ and $\kappa_D$ are scale and shape parameters respectively. $f_D$ is dependent only on an individual's age is given by

$$\frac{df_D}{da} = 1 - \frac{1-f_{D0}}{1+\left(\frac{a}{a_D}\right)^{\gamma_D}}$$

where $f_{D0}$ represents the time-scale at which immunity changes with age, and $a_D$ and $\gamma_D$ are scale and shape parameters respectively. Lastly, $\alpha_A$ and $\alpha_U$ are parameters that determine the probability that an individual in states A and U are detectable by PCR, which are given by $q^{\alpha_A}$ and $q^{\alpha_U}$ respectively.

The probability that an infected individual infects a mosquito upon being bitten is proportional to both their infectious state and their probability of detection, with a lower probability of detection assumed to correlate with a lower parasite density. Individuals who are in state D (clinically diseased), state U (sub-patent infection) and state T (receiving treatment) contribute to an onward infection within a mosquito with probabilities $c_D$, $c_U$ and $c_T$. In state *A*, contribution to an onward infection within a mosquito occurs with probability $c_A$, and is given by $c_U + \left(c_D - c_U\right)q^{\gamma_I}$ where $q$ is the probability of being detected by microscopy when asymptomatic, and $\gamma_I$ is a parameter that controls how quickly infectiousness falls within the asymptomatic state.

**Human Stochastic Model Equations**

Given the definitions above, the full stochastic individual-based human component of the model can be formally described by its Kolmogorov forward equations. As before, let $i$ index individuals in the population. Then, the state of individual $i$ at time $t$ is given by $\{j, k, t_k, l, t_l, m, t_m, a, t\}$, where $a$ is age, $j$ represents infection status ($S, D, A, U, T$ or $P$), $k$ is the level of infection-blocking immunity and $t_k$ is the time at which infection blocking immunity was last boosted. Similarly, $l$ and $t_l$ denote the level and time of last boosting of clinical immunity, respectively, while $m$ and $t_m$ do likewise for parasite detection immunity. Let $\delta_{p,q}$ denote the Kronecker delta ($\delta_{p,q} = 1$ if $p = q$ and 0 otherwise) and $\delta(x)$ denote the Dirac delta function. Defining $P_i(j, k, t_k, l, t_l, m, t_m, a, t)$ as the probability density function for individual $i$ being in state $\{j, k, t_k, l, t_l, m, t_m, a, t\}$ at time $t$, the time evolution of the system is governed by the following forward equation:

$$\frac{\partial P_i(j,k,t_k,l,t_l,m,t_m,a,t)}{\partial t} + \frac{\partial P_i(j,k,t_k,l,t_l,m,t_m,a,t)}{\partial a} =$$

$$\delta_{j,S}\left[r_P P_i(P, k, t_k, l, t_l, m, t_m, a, t) + r_U P_i(U, k, t_k, l, t_l, m, t_m, a, t)\right]$$

$$+ \delta_{j,A}\left[r_D P_i(D, k, t_k, l, t_l, m, t_m, a, t)\right]$$

$$+ \delta_{j,U}\left[r_A P_i(A, k, t_k, l, t_l, m, t_m, a, t)\right]$$

$$+ \delta_{j,P}\left[r_T P_i(T, k, t_k, l, t_l, m, t_m, a, t)\right]$$

$$+ (1 - b_i)\epsilon_i(t - d_E)\left[\delta_{j,S} + \delta_{j,D} + \delta_{j,A} + \delta_{j,U}\right] O_b \diamond P_i(j, k, t_k, l, t_l, m, t_m, a, t)$$

$$+ b_i\epsilon_i(t - d_E)\left[\delta_{j,A}(1 - \phi_i) + \delta_{j,D}\phi_i(1 - f_T) + \delta_{j,T}\phi_i f_T\right] O_b \diamond O_c \diamond O_d \diamond \sum_{j' \in \{S,A,U\}} P_i(j', k, t_k, l, t_l, m, t_m, a, t)$$

$$+ b_i h_i(t - d_E)\, O_b \diamond O_c \diamond O_d \diamond P_i(D, k, t_k, l, t_l, m, t_m, a, t)$$

$$+ \left[r_B k\frac{\partial}{\partial k} + r_{CA} l\frac{\partial}{\partial l} + r_{ID} m\frac{\partial}{\partial m}\right] P_i(j, k, t_k, l, t_l, m, t_m, a, t)$$

$$+ \mu\, \delta(a)\delta(t_k + T_{big})\delta(t_l + T_{big})\delta(t_m + T_{big})\delta_{j,S}\delta_{k,0}\delta_{l,0}\delta_{m,0}\sum_{j'} P_i(j', k, t_k, l, t_l, m, t_m, a, t)$$

$$-\left[\mu + r_P\delta_{j,P} + r_U\delta_{j,U} + r_D\delta_{j,D} + r_A\delta_{j,A} + r_T\delta_{j,P} + h_i(t - d_E)\left[\delta_{j,S} + \delta_{j,D} + \delta_{j,A} + \delta_{j,U}\right]\right] P_i(j, k, t_k, l, t_l, m, t_m, a, t)$$

Here $O_b$, $O_c$ and $O_d$ are commutative integral operators with the following action on a density $(j, k, t_k, l, t_l, m, t_m, a, t)$ :

$$O_b \diamond f = \delta(t - t_k) \int_0^\infty f(j, k - 1, t - u_B - \tau, l, t_l, m, t_m, a, t) d\tau + \theta\left(\frac{t - t_k}{u_B}\right) f(j, k, t_k, l, t_l, m, t_m, a, t)$$

$$O_c \diamond f = \delta(t - t_l) \int_0^\infty f(j, k, t_k, l - 1, t - u_C - \tau, m, t_m, a, t) d\tau + \theta\left(\frac{t - t_l}{u_C}\right) f(j, k, t_k, l, t_l, m, t_m, a, t)$$

$$O_d \diamond f = \delta(t - t_m) \int_0^\infty f(j, k, t_k, l, t_l, m - 1, t - u_D - \tau, a, t) d\tau + \theta\left(\frac{t - t_m}{u_D}\right) f(j, k, t_k, l, t_l, m, t_m, a, t).$$

Finally, $\theta(x)$ is an indicator function such that $\theta(x) = 1$ if $x < 1$ and 0 otherwise.

For simulation, a discrete time approximation of this stochastic model was used, with a time-step of 1 day. For each individual $k$, $l$ and $m$ are set to zero at birth, while $t_k$, $t_l$ and $t_m$ are set to a large negative value $-T_{big}$ (to represent never having been exposed or infected, i.e. their immunity will always be boosted upon their first exposure or infection event). Each immunity term increases by 1 for an individual whenever that individual receives an infectious bite ($k$), or is infected ($l$ and $m$), if the previous boost to $k$, $l$ and $m$ occurred more than $u_B$, $u_C$ and $u_D$ days earlier, respectively. Immunity levels decay exponentially at rate $r_B$, $r_{CA}$ and $r_{ID}$, where $r_B$, $r_{CA}$ and $r_{ID}$ are equal to $\frac{1}{d_B}$, $\frac{1}{d_{CA}}$ and $\frac{1}{d_{ID}}$ respectively.

#### <u>Mosquito Transmission Model</u>

##### Mosquito Population Dynamics

The adult stage of mosquito development was modelled individually and is similarly described in its deterministic framework before exploring its stochastic implementation. Adult mosquitoes will begin life susceptible to infection ($S_M$), and will seek a blood meal on the same day they are born and every 3 days after that until the mosquito dies. Each feeding day, mosquito *i* will be exposed to a force of infection, $\Lambda_{Mi}$, depending on the infection status and immunity of the human the mosquito is feeding on. The overall force of infection towards the mosquito population on a given day, $\Lambda_M$, is thus represented by the sum of the onward infection contributions from each infected human, delayed by $d_g$, delay due gametocytogenesis, which is given by

$$\Lambda_M = \alpha_k Q_0 \left( \sum_{i=1}^{\Sigma_D} \pi_i c_D + \sum_{i=1}^{\Sigma_T} \pi_i c_T + \sum_{i=1}^{\Sigma_A} \pi_i c_A + \sum_{i=1}^{\Sigma_U} \pi_i c_U \right) (t - d_g)$$

where $\alpha_k$ is the daily rate at which a mosquito takes a blood meal, $Q_0$ is the proportion of bites that are on humans (anthropophagy) and $d_g$ represents the delay from emergence of asexual blood-stage parasites to sexual gametocytes that contribute towards onward infectivity. Infected mosquitoes then pass through a latent infection stage ($E_M$) that will last 10 days, representing the extrinsic incubation period for the parasite ($d_{EM}$), before becoming infectious to humans ($I_M$). Infectious mosquitoes remain infectious until they die. Whenever a mosquito dies, it is replaced with a new susceptible adult mosquito. Analogously to the human model, when a new adult mosquito emerges, the day on which it dies is drawn from an exponential distribution with a transition rate of $\mu_M$= 0.132 days. The differential equations summarising the adult stage of mosquitoes are given by

$$\frac{dS_M}{dt} = \mu_M M_v \ - \mu_M S_M \ - \Lambda_M S_M$$

$$\frac{dE_M}{dt} = \Lambda_M S_M - \mu_M E_M \ - \Lambda_M(t - d_{EM}) S_M(t - d_{EM}) exp^{-\mu_M d_{EM}}$$

$$\frac{dI_M}{dt} = \Lambda_M(t - d_{EM}) S_M(t - d_{EM}) exp^{-\mu_M d_{EM}}$$

where $\mu_M$ is the daily death rate of adult mosquitoes, and $M_v$ is the total mosquito population, i.e. $S_M + E_M + I_M$.

**Mosquito Stochastic Model Equations**

As with the human transmission model, the full stochastic individual-based mosquito component of the model can be formally described by its Kolmogorov forward equations. As before, let $i$ denote each mosquito in the population, and $j$ denote their infection status. Let $\delta_{p,q}$ denote the Kronecker delta function such that it equals 1 if $p = q$ and 0 otherwise. Defining $P_i(j, t)$ as the probability density function for mosquito $i$ being in state $\{j, t\}$ at time $t$, the time evolution of the system is governed by the following forward equation:

$$\frac{\partial P_i(j, t)}{\partial t} = \ \delta_{j,E_M}\left[\Lambda_{Mi}\left(P_i\left(S_M, t\right)\right)\right] + \ \delta_{j,I_M}\left[\Lambda_{Mi}\left(t - d_{EM}\right)\left(P_i\left(S_M, t\right)\right)\right] +$$

$$\delta_{j,S_M}\mu_M\left[P_i\left(S_M, t\right) + P_i\left(E_M, t\right) + \ P_i\left(I_M, t\right)\right] - \ P_i(j, t)\left[\mu_M \ + \Lambda_{Mi}\left[\delta_{j,S_M}\right] \ + \Lambda_{Mi}\left(t - d_{EM}\right)\left[\delta_{j,S_M}\right]\right]$$

#### Parasite Dynamics

##### Parasite Genetic Barcode

Parasites are modelled as discrete populations as a result of an infection event associated with a mosquito or a human. Each asexual parasite is characterised by one genetic barcode, which encodes for biallelic single nucleotide polymorphisms (SNPs) distributed across the parasite genome. Sexual stages of the parasite lifecycle within the mosquito are represented by both a female and male barcode, thus defining the range of recombinants that could be produced. The within human parasite dynamics and model considerations are discussed first before exploring the within mosquito parasite life cycle and associated modelling implications. A schematic overview of the modelled parasite lifecycle stages is shown in Supplementary Figure X.

##### Within Human Parasite Dynamics

During a successful mosquito to human infection event, at least one asexual parasite barcode is introduced into the human, which will be observed in the ensuing gametocyte genotypes when considering onward infectiousness from humans to mosquitoes. More than one different asexual parasite barcode that will be observed in the ensuing gametocyte genotypes may be introduced during an infection event, representing cotransmission of genetically related parasites (assuming the mosquito was infected with more than one sporozoite genotype). The precise distribution describing the number of genotypes is unknown,[9] but the mean number of sporozoites within an innoculation event is well characterised by a geometric distribution with the mean equal to 10. The geometric mean will then be used to estimate the proportion of sporozoites that are successful, ξ, which yields the maximum number of successful sporozoites in an individual with no pre-erythrocytic immunity. The observed number of successful sporozoites is then calculated by conducting Bernoulli trials for all but one of the successful sporozoites (as we assume one has to survive to found the infection) to see if they are successful, calculate using the individual's probability of infection, $b_i$. In summary, this can be written as:

$$Total_{spz} \sim Geom\left(p_{spz} = 0.1\right)$$

$$Max_{spz} = ceiling\left(Total_{spz}\xi\right)$$

$$Observed_{spz} = 1 + \sum_{1}^{Max_{spz}-1} bernoulli(b_i)$$

There is no assumed maximum number of parasites, with individuals assumed to clear strains on the day that they would have moved from a subpatent infection to susceptible for the strain considered, i.e. each acquired strain follows an assumed trajectory in parasitaemia representative of a normal infection cycle, i.e. with a mean duration of infectiousness equal to $d_A + d_U$. Acquired strains can thus move "infection state" independently of the human's infection state. For example, an individual is infected on day 0 and develops an asymptomatic infection. The individual would have moved to a subpatent stage on day 200, but they were bitten on day 150, and developed clinical symptoms, moving to state D. When this happens, the strain acquired on day 0 is still considered to be at the parasitaemia if the individual had not been infected again. Consequently, this strain will become a "subpatent" strain on day 200. By tracking parasites in this way, we are able to track the relative parasitemias of each acquired strain, enabling more accurate sampling of within host parasite genetic diversity when passing on gametocytes to mosquitoes. This is shown below (**Supplementary Methods Figure 2)**, which also details the key features of the SNP barcode.

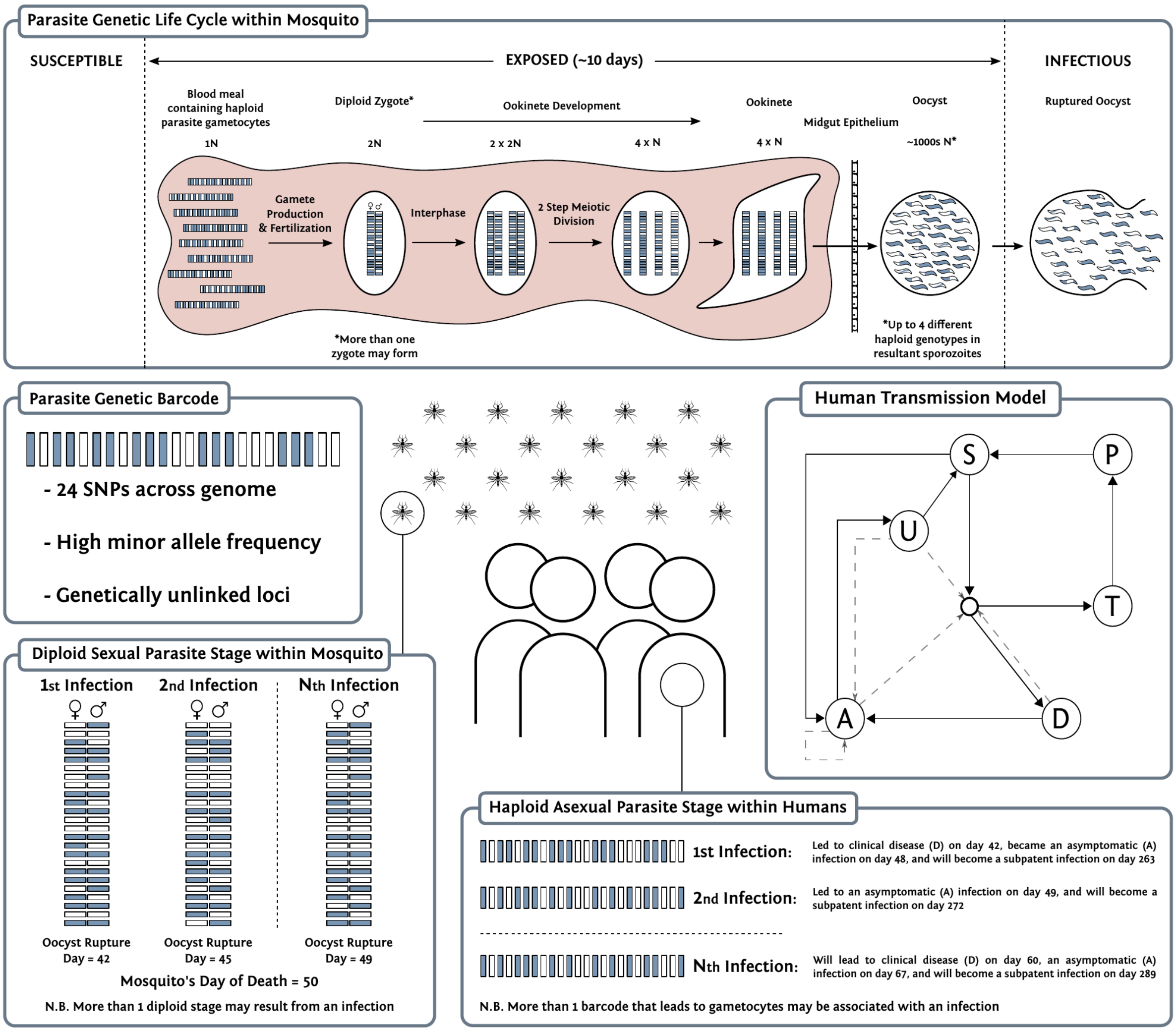


**Supplementary Methods Figure 2: Parasite dynamics within the transmission model.** Individual mosquitoes are tracked, which allows for recombination to be modelled explicitly. Populations of parasite clones are tracked, and multiple oocysts are able to be formed from a feeding event, as well as multiple genetically distinct sporozoites onwardly transmitted. A "barcode" is associated with each parasite clone and encodes for specific biallelic SNPs in the parasite genome.

**Within mosquito parasite dynamics**

When a mosquito is infected, we sample from a zero-truncated negative binomial distribution that describes the distribution of oocysts that form from a feeding event. The choice of a zero truncated negative binomial represents the increasingly identified zero-inflated negative binomial that describes the relationship between oocyst prevalence and mean oocysts per mosquito in SMFA studies.[22–24] The related negative binomial distribution for the distribution of oocysts is given by

$$X_{oocysts} \sim NB(size_{oocysts}, shape_{oocysts})$$

where $X_{oocysts}$ represents the number of oocysts that will be formed, with mean and shape equal to 5.[36–38] For each oocyst formed, two barcodes are sampled from the infected host representing the female and male gametes that led to the oocysts formation. These two barcodes will result in up to 4 different potential genotypes (reflecting the immediate two step meiotic division that takes place after zygote formation) represented within the sporozoite population within the oocyst. When an infectious mosquito seeks a blood meal and leads to an onward infection, a value for $Observed_{spz}$ is sampled. The oocyst source for each onward infection within a coinfection is sampled from oocysts that have ruptured, i.e. the infection event that led to the oocyst occurred more than 10 days earlier. At this point, recombination is simulated by randomly choosing either the male or female allele at each SNP position in the barcode. The random sampling in this represents the assumed independent segregation events resulting from the absence of genetic linkage between barcode SNP positions. Once a recombinant has been simulated, it is stored and associated with the oocyst from which it came. If the same oocyst is chosen to lead to an additional infection, then the previously generated recombinant has a 25% chance of being onwardly transmitted and there is a 75% chance that a new recombinant is generated and subsequently saved. This process will continue in ensuing onward infection events that result from this oocyst until four recombinants have been simulated, at which point they each have a 25% chance of being onwardly transmitted. The above thus introduces an assumption that sporozoites will remain onwardly-transmissible for the remainder of the mosquito's life, with no effect upon their relative probability of being onwardly transmitted in relation to sporozoites that resulted from a more recently ruptured oocyst.

Full transmission model parameters are given in **Supplementary Methods Table 1**

#### Transmission Model Parameter Values

**Supplementary Methods Table 1:** Parameter estimates used within the model were taken from Griffin et al. 2014,[4] 2015[3] and 2016[2] and Watson et al. 2022.

| Parameter | Symbol | Estimate |
|---|---|---|
| Human infection duration (days) | | |
| Latent period | $d_E$ | 12 |
| Patent infection | $d_A$ | 200 |
| Clinical disease (treated) | $d_T$ | 5 |
| Clinical disease (untreated) | $d_D$ | 5 |
| Sub-patent infection | $d_U$ | 110 |
| Prophylaxis following treatment | $d_P$ | 25 |
| Treatment and Importation Parameters | | |
| Probability of seeking treatment if clinically diseased | $f_T$ | Variable |
| Importation Rate | $\delta_{imports}$ | 0.01 |
| Infectiousness to mosquitoes | | |
| Lag from parasites to infectious gametocytes | $d_g$ | 12 days |
| Untreated disease | $c_D$ | 0.0680 day$^{-1}$ |
| Treated disease | $c_T$ | 0.0219 day$^{-1}$ |
| Sub-patent infection | $c_U$ | 0.000620 day$^{-1}$ |
| Parameter for infectiousness of state A | $\gamma_1$ | 1.824 |
| Age and heterogeneity | | |
| Age-dependent biting parameter | $\rho$ | 0.85 |
| Age-dependent biting parameter | $a_0$ | 8 years |
| Daily mortality rate of humans | $\mu$ | 0.000180 |
| Variance of the log heterogeneity in biting rates | $\sigma^2$ | 1.67 |
| Immunity reducing probability of infection | | |
| Maximum probability due to no immunity | $b_0$ | 0.590 |
| Maximum relative reduction due to immunity | $b_1$ | 0.5 |
| Inverse of decay rate | $d_B$ | 10 years |
| Scale parameter | $I_{B0}$ | 43.879 |
| Shape parameter | $\kappa_B$ | 2.155 |
| Duration in which immunity is not boosted | $u_B$ | 7.199 |

| Immunity reducing probability of clinical disease | | |
|---|---|---|
| Maximum probability due to no immunity | $\phi_0$ | 0.791 |
| Maximum relative reduction due to immunity | $\phi_1$ | 0.000737 |
| Inverse of decay rate | $d_{CA}$ | 30 years |
| Scale parameter | $I_{C0}$ | 18.0237 |
| Shape parameter | $\kappa_C$ | 2.370 |
| Duration in which immunity is not boosted | $u_C$ | 6.0635 |
| New-born immunity relative to mother's | $P_M$ | 0.774 |
| Inverse of decay rate of maternal immunity | $d_M$ | 67.695 |
| Immunity reducing probability of detection | | |
| Minimum probability due to maximum immunity | $d_1$ | 0.161 |
| Inverse of decay rate | $d_{ID}$ | 10 years |
| Scale parameter | $I_{D0}$ | 1.578 |
| Shape parameter | $\kappa_D$ | 0.477 |
| Duration in which immunity is not boosted | $u_D$ | 9.445 |
| Scale parameter relating age to immunity | $a_D$ | 21.9 years |
| Time-scale at which immunity changes with age | $f_{D0}$ | 0.00706 |
| Shape parameter relating age to immunity | $\gamma_D$ | 4.818 |
| PCR detection probability parameters state A | $\alpha_A$ | 0.757 |
| PCR detection probability parameters state U | $\alpha_U$ | 0.186 |
| Mosquito Population Model | | |
| Daily mortality of adults | $\mu_M$ | 0.132 |
| Daily biting rate | $\alpha_k$ | 0.333 |
| Anthropophagy | $Q_0$ | 0.92 |
| Extrinsic incubation period | $d_{EM}$ | 10 days |
| Negative Binomial shape parameter for distribution of oocyst frequencies upon infection | $shape_{oocysts}$ | 5 |
| Negative Binomial size parameter for distribution of oocyst frequencies upon infection | $size_{oocysts}$ | 5 |
| Human Parasite Parameters | | |
| Geometric distribution of total sporozoites in an infectious bite probability | $p_{spz}$ | 0.1 |
| Proportion of successful sporozoites | $\xi$ | 22% |

#### Parasite genetic barcode for modelling resistance

In order to model antimalarial resistance, we have adapted the parasite barcode used in Watson et al. for the simulation of resistance.[5] Each position in the barcode represents either the absence (barcode position is equal to 0) or presence (barcode position is equal to 1) of a resistance mutation associated with resistance to a particular drug. For example, **Supplementary Methods Table 2** shows how a barcode with three loci can be used to represent resistance to DHA-PPQ and ASMQ.

**Supplementary Methods Table 2.** Example barcode alterations to model antimalarial resistance

| Artemisinin Resistance | Piperaquine Resistance | Mefloquine Resistance | Phenotype |
|---|---|---|---|
| 0 | 0 | 0 | Wild type parasite. Fully susceptible. |
| 1 | 0 | 0 | Resistant to artemisinin |
| 0 | 1 | 0 | Resistant to PPQ |
| 0 | 0 | 1 | Resistant to MQ |
| 1 | 1 | 0 | Resistant to DHA-PPQ |
| 1 | 0 | 1 | Resistant to ASMQ |
| 0 | 1 | 1 | Resistant to PPQ and MQ |
| 1 | 1 | 1 | Multidrug resistant to DHA-PPQ and ASMQ |

Barcodes are used to track populations of parasites that are introduced from an infectious mosquito bite. We assume loci are genetically unlinked (genes known to confer resistance to the five first-line ACTs recommended by the WHO each occurring on different chromosomes) and consequently segregate independently during recombination.

**Fitness Costs**

Antimalarial resistance is assumed to introduce a fitness cost to resistant parasites compared to wild-type parasites. Fitness costs are associated with each resistance locus and are assumed to be multiplicative. The fitness cost manifests as a reduction in parasite density that is assumed to reduce the probability of the resistant parasite being passed on to a mosquito. This can be expressed mathematically as follows. Let $b = \left[b_1, b_2, ..., b_m\right]$ describe the vector of barcode loci for a barcode of length $m$. For example, the ASMQ resistant strain in **Supplementary Methods Table 2** is represented by the vector $[1, 0, 1]$. If $\nu_j$ is the resistance cost associated with barcode locus $j$, then the comparative fitness cost due to resistance, $r$, for the given parasite is simply:

$$r = \prod_{i=j}^{m}(1 - b_j v_j)$$

Consequently, in this study, the wild-type allele at each locus is assumed to confer no fitness costs, with $(1 - b_j v_j)$ equal to 1 if locus $j$ is wild-type, and thus $r = 1$ for the wild-type parasite in **Supplementary Methods Table 2**. In this analysis, each resistance locus is assumed to have a fitness cost equal to 0.0005, i.e. $v_{i...m} = 0.0005$. As a result, the relative fitness of a genotype with $k$ resistance mutations, in the absence of any drugs present in the blood, is $(1 - 0.005)^k$.

The number of oocysts generated from each mosquito bite on an infectious human is drawn from a zero-truncated negative binomial distribution with mean = 2.5 and shape = 1. The selection of which parasite strains from the infected individual contributed to the oocysts is given by the relative probability that a given strain will be chosen in an individual with $n$ gametocytogenic parasite strains, and is given by:

$$cr = \left[c_1 r_1, c_2 r_2, ... c_n r_n\right]$$

$r_i$ is the fitness cost associated with parasite strain $i$ and $c_i$ is the contribution of parasite $i$ to onward infection, which will be either $c_T$, $c_D$, $c_A$ or $c_U$ depending on the infection status of strain $i$ denoted here as $X_i$. As in the original model, $c_T$, $c_D$ and $c_U$ are constants, and $c_A$ is dependent on an individual's immunity. The probability that an infected individual infects a mosquito is still determined by the set of parameters determining the onward contribution to transmission, $\left\{c_T, c_D, c_A, c_U\right\}$, which are based on the infection status and immunity of the individual, denoted here as $Y$. However, if the strains responsible for the human's current infection state, i.e. all strains that match the human's infection state, are resistant then the probability of onward transmission is determined by the highest onward contribution of these strains, which is given by:

$$max\left\{cr_i : X_i = Y\right\}$$

In this way, fitness costs both affect the relative probability that a resistant strain is transmitted compared to a wild-type strain in a mixed infection, while also reducing the probability that transmission occurs in individuals where the highest parasite density strain is resistant.

**Clinical and Treatment Outcomes**

With the addition of resistance, the treatment efficacy now varies and is determined both by the genotype of the parasite strains, the parasite density of each strain and the drug used to treat the infection. The probability that any given strain is cleared by drug $z$ can be expressed as $ez_{\beta(b)}$, where $\beta(b)$ is an adapted conversion of binary to decimal integers and is given by:

$$\beta(b) = \left(\sum_{j=1}^{m} b_j . 2^{j-1}\right) - 1$$

Using the same representation of a parasite genotype as in **Supplementary Methods Table 2**, the efficacy of drug $z$ can be expressed by the vector $ez = \left[ez_1, ez_2, ..., ez_{2^m}\right]$ for simulations in which the number of loci being modelled is equal to $m$. $ez_1$ represents the efficacy of the drug against the wild-type parasite, i.e. the barcode vector $b$ represented by a vector of length $m$ filled with zeros. Importantly, $ez$ reflects the probability that the drug will clear a parasite that has led to a symptomatic infection, i.e. the parasite strain is at a sufficiently high parasite density to trigger symptoms and seek treatment. This is equivalent to the probability of successfully clearing a symptomatic infection such that the infection does not recrudesce and lead to a 28-day treatment failure.

In this analysis, we used the drug efficacy by genotype table parameterised in [6], which was used by each group to parameterise the efficacy of each drug on each resistance genotype. 64 genotypes are included by allowing for variation at the K76T locus in *pfcrt*, the N86Y and Y184F loci in *pfmdr1*, the C580Y locus in pfkelch13, copy-number variation (CNV) of *pfmdr1*, and CNV of the *plasmepsin-2,3* genes. CNV is only separated into 'single copy' or 'multiple copies'. See Section 3 - Treatment efficacy on specific genotypes for more information ~~about this~~. In the Imperial model, the efficacy of each drug determines the probability of a 28-day parasitological failure, with the probability of late parasitological failure for each parasite genotype modelled shown in **Supplementary Methods Table 3**.

Parasites from previous infections are assumed to be at a lower parasite density than the infecting strain that triggered the clinical infection (and hence why the individual is seeking treatment) and will be more likely to be cleared by the drug. In the model, the infection state of each strain, $X_i$, as well as the day the strain was acquired, $t_0$, and the time the strain will move out of the infection state, $t_1$, is tracked. This information is used to define the probability that strain $i$ will recrudesce after treatment, $P(Recrudesce)_i$, which is given by:

$$P(Recrudesce)_i = \begin{cases} ez_{\beta(b_i)}\,, & X_i = Y \\ ez_{\beta(b_i)}\left(\frac{t_1 - t_c}{t_1 - t_0}\right), & X_i = A \\ 0\,, & X_i = U \end{cases}$$

where $t_c$ is the current time. $P(Recrudesce)_i$ assumes that parasites below ~200p/µl (state U) will always be cleared regardless of the parasite phenotype. $P(Recrudesce)_i$ also assumes that the probability that an asymptomatic parasite above ~200p/µl (state A) will recrudesce is linearly related to the age of the infection and is at its highest when it first enters state A.

The probability that a treated infection will be successfully cleared by drug $z$, $P(Cleared)$, is equal to one minus the highest probability of a strain recrudescing, which is given by:

$$P(Cleared) = 1 - max\,\{P(Recrudesce)_i : 0 \leq i \leq n\}$$

This assumes that the multiplicity of infection does not directly affect the probability that an individual will be cleared, i.e. only one Bernoulli trial with probability $P(Cleared)$ is used to determine if all parasite strains were cleared.

If an individual fails treatment, it is assumed that they will recrudesce to yield a late parasitological failure (LPF) and move into state A after the prophylactic period of the drug has finished. Whether each parasite strain in a multiply infected individual recrudesces during a LPF is dependent on $P(Recrudesce)_i$. Bernoulli trials are conducted for each parasite strain except for one random strain for which $P(Recrudesce)_i = P(Cleared)$, which ensures that one of the most likely strains to recrudesce did actually recrudesce and cause a LPF.

If an individual successfully clears all parasites, they will either move directly into a state P or they will remain in the treated compartment for a longer duration, resulting from slow parasite clearance (SPC). SPC is assumed to always occur if any of the pre-treatment strains that contributed to the clinical disease were resistant to any component of the drug. The duration of SPC was set equal to 6 days based on previous modelling studies estimating parasite clearance rates associated with SPC for both AL and ASAQ, and a longer duration of 9 days for DHA-PPQ.[7] During SPC it is assumed that all parasite strains not resistant to the drug given have cleared and will thus not contribute to onward infection during SPC.

Lastly, individuals in state P can be reinfected before returning to state S, depending on the genotype of the infecting strain, how recently they were treated and the ACT used, reflecting the different half-lives of available partner drugs. For all drugs, the probability of reinfection in state P increases as the partner drug wanes. As described in [8], we use the gamma distribution, $\Gamma(\alpha_i, \beta_i)$, where $\alpha_i$ and $\beta_i$ are the shape and rate parameter respectively of the Gamma distribution, for drug $i$, to describe the probability of reinfection in individuals treated with AL and ASAQ. We use $\alpha_{AL} = 93.5$ and $\alpha_{ASAQ} = 16.8$ for the shape parameters for AL and ASAQ. These values represent the mean posterior estimate for the shape parameter in [8], which were estimates across a number of trial sites in Africa. We use $\beta_{AL} = 5.22$ and $\beta_{ASAQ} = 0.94$ for the rate parameters, which yield mean durations of protection equal to 17.9 days and 17.8 days, respectively. These durations are longer than the posterior mean duration of protection estimated in [8] and more closely reflect study sites in [8] with the longest times to reinfection for AL and ASAQ. This was chosen because the mean duration was estimated across study sites with known resistance markers and subsequently we chose to match to relationships in sites that were assumed to have the least amount of partner drug resistance for each drug. For DHA-PPQ, we assume the probability of reinfection is described by a Weibull survival curve, with scale and shape equal to 28.1 and 4.4 respectively, as estimated in [9] , with a mean duration of protection equal to 25 days. The described periods of prophylaxis are used to define the per-day probability of an individual being reinfected if the parasites introduced during an infectious bite are not resistant to the partner drug. However, if any of the introduced parasites are resistant to the partner drug, we assume a shorter period of prophylaxis. For AL and ASAQ, we use $\beta_{AL} = 10.75$ and $\beta_{ASAQ} = 1.45$, resulting in mean durations of protection equal to 8.7 days and 11.6 days respectively. These shorter durations were chosen to match ~~to~~ prophylactic profiles in sites in [8] with the shortest durations of protection due to the presence of partner drug resistance. For DHA-PPQ, we used a larger scale parameter of 59 for the Weibull survival curve, approximately halving the mean duration of protection to 12.4 days.

In order to simulate individual level variation in drug clearance and prophylaxis we first fit a Hill function to relate the assumed exponential clearance of the partner drug to the described period of prophylaxis for each drug on both wild-type and resistant parasites. For each drug, we assume the mean drug lifetime is equal to $ln(2)$ * mean duration of prophylaxis of each drug against wild-type parasites (17.9, 17.8 and 25 days for AL, ASAQ and DHA-PPQ respectively from earlier prophylaxis fitting). When an individual moves from being treated to being in a state of prophylaxis, we draw the time at which they will become fully susceptible again from an exponential distribution with a mean equal to the mean duration of prophylaxis for each drug. During this prophylactic period, we assume the individual's drug concentration decays exponentially such that they have fully eliminated the partner drug when returning to state S, with their drug concentration on each day used to calculate the probability of early reinfection using the earlier fitted Hill functions.

**Supplementary Methods Table 3.** Parasite Genotype to 28-day Treatment Failure Phenotype. Sourced from Watson et al. 2022.[1]

| Genotype | DHAPPQ | ASAQ | AL |
|---|---|---|---|
| KNY1C1 | 0.972486 | 0.962073 | 0.915158 |
| TNY1C1 | 0.972486 | 0.947158 | 0.929358 |
| KYY1C1 | 0.972486 | 0.905281 | 0.953453 |
| TYY1C1 | 0.972486 | 0.891124 | 0.964582 |
| KNF1C1 | 0.972486 | 0.976982 | 0.889945 |
| TNF1C1 | 0.972486 | 0.958879 | 0.908054 |
| KYF1C1 | 0.972486 | 0.930465 | 0.915158 |
| TYF1C1 | 0.972486 | 0.917383 | 0.929358 |
| KNY2C1 | 0.972486 | 0.962073 | 0.858645 |
| TNY2C1 | 0.972486 | 0.947158 | 0.869483 |
| KYY2C1 | 0.972486 | 0.905281 | 0.896817 |
| TYY2C1 | 0.972486 | 0.891124 | 0.915158 |
| KNF2C1 | 0.972486 | 0.976982 | 0.830015 |
| TNF2C1 | 0.972486 | 0.958879 | 0.844322 |
| KYF2C1 | 0.972486 | 0.930465 | 0.858645 |
| TYF2C1 | 0.972486 | 0.917383 | 0.869483 |
| KNY1Y1 | 0.928664 | 0.895737 | 0.795336 |
| TNY1Y1 | 0.928664 | 0.864359 | 0.828863 |
| KYY1Y1 | 0.928664 | 0.771709 | 0.877727 |
| TYY1Y1 | 0.928664 | 0.735268 | 0.907645 |
| KNF1Y1 | 0.928664 | 0.948359 | 0.723351 |
| TNF1Y1 | 0.928664 | 0.892625 | 0.753201 |
| KYF1Y1 | 0.928664 | 0.824725 | 0.795336 |
| TYF1Y1 | 0.928664 | 0.794297 | 0.828863 |
| KNY2Y1 | 0.928664 | 0.895737 | 0.646288 |
| TNY2Y1 | 0.928664 | 0.864359 | 0.684442 |
| KYY2Y1 | 0.928664 | 0.771709 | 0.75155 |
| TYY2Y1 | 0.928664 | 0.735268 | 0.795336 |
| KNF2Y1 | 0.928664 | 0.948359 | 0.570356 |
| TNF2Y1 | 0.928664 | 0.892625 | 0.604481 |
| KYF2Y1 | 0.928664 | 0.824725 | 0.646288 |
| TYF2Y1 | 0.928664 | 0.794297 | 0.684442 |
| KNY1C2 | 0.768484 | 0.962073 | 0.915158 |
| TNY1C2 | 0.768484 | 0.947158 | 0.929358 |
| KYY1C2 | 0.768484 | 0.905281 | 0.953453 |
| TYY1C2 | 0.768484 | 0.891124 | 0.964582 |
| KNF1C2 | 0.768484 | 0.976982 | 0.889945 |
| TNF1C2 | 0.768484 | 0.958879 | 0.908054 |
| KYF1C2 | 0.768484 | 0.930465 | 0.915158 |
| TYF1C2 | 0.768484 | 0.917383 | 0.929358 |
| KNY2C2 | 0.768484 | 0.962073 | 0.858645 |
| TNY2C2 | 0.768484 | 0.947158 | 0.869483 |
| KYY2C2 | 0.768484 | 0.905281 | 0.896817 |
| TYY2C2 | 0.768484 | 0.891124 | 0.915158 |
| KNF2C2 | 0.768484 | 0.976982 | 0.830015 |
| TNF2C2 | 0.768484 | 0.958879 | 0.844322 |
| KYF2C2 | 0.768484 | 0.930465 | 0.858645 |
| TYF2C2 | 0.768484 | 0.917383 | 0.869483 |
| KNY1Y2 | 0.414973 | 0.895737 | 0.795336 |
| TNY1Y2 | 0.414973 | 0.864359 | 0.828863 |
| KYY1Y2 | 0.414973 | 0.771709 | 0.877727 |
| TYY1Y2 | 0.414973 | 0.735268 | 0.907645 |
| KNF1Y2 | 0.414973 | 0.948359 | 0.723351 |
| TNF1Y2 | 0.414973 | 0.892625 | 0.753201 |
| KYF1Y2 | 0.414973 | 0.824725 | 0.795336 |
| TYF1Y2 | 0.414973 | 0.794297 | 0.828863 |
| KNY2Y2 | 0.414973 | 0.895737 | 0.646288 |
| TNY2Y2 | 0.414973 | 0.864359 | 0.684442 |
| KYY2Y2 | 0.414973 | 0.771709 | 0.75155 |
| TYY2Y2 | 0.414973 | 0.735268 | 0.795336 |
| KNF2Y2 | 0.414973 | 0.948359 | 0.570356 |
| TNF2Y2 | 0.414973 | 0.892625 | 0.604481 |
| KYF2Y2 | 0.414973 | 0.824725 | 0.646288 |
| TYF2Y2 | 0.414973 | 0.794297 | 0.684442 |

#### Mapping antimalarial resistance

##### Prevalence Data Assembly and Aggregation

To systematically evaluate the prevalence and distribution of antimalarial drug resistance across Africa, our study integrated data from three key databases: the WHO Threat Maps, the WorldWide Antimalarial Resistance Network, and the Pf7k database. These sources provided comprehensive data on resistance markers, including the K76T locus in the pfcrt gene, N86Y and Y184F in the pfmdr1 gene, C580Y in pfkelch13, as well as copy number variations (CNV) in the pfmdr1 and the plasmepsin 2 and 3 genes of Plasmodium falciparum. We treated CNV as a binary variable to distinguish between single and multiple copies, resulting in a profile of 64 potential genotypes to track, which are crucial for linking genotypic profiles to resistance phenotypes and inferring treatment outcomes with antimalarials like AL, ASAQ, and DHAPPQ.

In brief, all surveys assessed P. falciparum samples collected between 2004 and 2018 in sub-Saharan Africa for genotypes pfmdr1 N86Y, Y184F, and D1246Y and/or pfcrt K76T. We excluded studies that reported prevalence data aggregated across multiple sites more than 300 km apart and whose results could not be disaggregated (n = 23 surveys) (35). From each survey, we extracted the midpoint year of sampling, geographic coordinates of the study site, the number of samples tested at each locus, and the number of samples with wild-type, mutant, or mixed genotypes (in the case of polygenomic infections) at each locus. Prevalence was calculated as the number of individuals with mutant or mixed infections out of the total surveyed (35). The raw data underwent rigorous preprocessing, including cleaning, deduplication, removal of mixed infection data, and filtering to retain only those studies with precise sample locations and collection dates. This process yielded a refined dataset of 7,390 samples from 1998 to 2021, representing resistance frequency data.

While we define prevalence as the number of individuals with mutant or mixed infections out of the total number of P. falciparum–infected individuals surveyed, frequency is the proportion of parasite clones with the molecular marker in the parasite population, which may matter in highly endemic regions in which individuals often have polygenomic infections (35, 45). For example, say an individual is infected with four clones of P. falciparum, one of which has the mutant genotype of interest: If only that single person were surveyed, the frequency of the mutation would be 25%, whereas the prevalence would be 100%. Because the majority of studies (n = 209/254 studies) did not report the average number of clones within the surveyed population, we used the subset of data that reported the prevalence of mixed and mutant infections separately (n = 245/501 surveys) and inferred the proportion of multiclonal infections using the P. falciparum parasite rate obtained from the Malaria Atlas Project (44). We then applied methodology developed by Okell et al. (35) to estimate the observed frequency of each marker in each survey and aggregated surveys in the same method as described above.

To visualise the spatial distribution of current resistance prevalence, we adopted the methodology used by Ehrlich et al. (2021) for estimating the frequency of sulfadoxine-pyrimethamine (SP) resistance. This involved calculating the weighted average prevalence of resistance markers within each first-level administrative region in Africa over distinct study periods (1998-2004, 2004-2010, 2010-2016, and 2016-2021). The choice of this methodological approach was necessitated by the spatiotemporal sparsity of the data, which precluded the use of continuous geostatistical models.

**Covariate Data Assembly and Selection**

To statistically model resistance frequency, we included variables previously hypothesised to be associated with drug resistance.  This allowed for improved predictions in regions lacking direct observational data. We selected covariates hypothesised to influence drug resistance, including malaria prevalence, treatment-seeking rates, and average travel times to healthcare facilities at the first administrative level, with the proportion of artemisinin-based combination therapy (ACT) used and GDP per capita included at the national level. For all covariates, excluding those only available at the national level, we averaged pixel values from rasterized data to generate administrative division-level estimates. Covariates were extracted for the midpoint year of both time periods, when possible, and for the equivalent study region.

**Bayesian Hierarchical Spatial Model**

We used a multivariate logistic regression framework with spatially correlated random effects to identify strongly predictive covariates that explain variability in the frequency of resistance genotypes observed across first-administrative regions, which could be used to predict resistance frequency in regions and times without observed data. We previously found evidence of spatial autocorrelation in the data, suggesting utility of spatial analysis (14). Here, we specify a conditional autoregressive (CAR) model for the random effects which allows for the possibility that neighbouring administrative divisions exhibit similar behaviour, leading to localised smoothness of estimated prevalence, potentially improved predictions in unobserved regions, and accurate statistical inference for the regression associations of interest. The model for the aggregated survey data was fit during selected time periods with the final time period between 2017-2021, which was used to infer the starting resistance frequency in forward simulations.

Each resistance marker and time period was modelled separately using Bayesian inference techniques. Markov chain Monte Carlo (MCMC) sampling was employed with four chains of 110,000 iterations, with a burn-in phase of 10,000 and thinned by a factor of 100. Weakly informative priors to guide parameter estimation were used and after ensuring convergence through visual inspection of trace plots and calculation of the Geweke diagnostic for all monitored parameters and assessing the quality of model fit, 1000 draws were sampled from the posterior distribution to construct median estimates and 95% confidence intervals for the frequency of each resistance marker. Spatial models were implemented with the CARBayes package (48).

#### Modelling the spread of artemisinin resistance

We trained an ensemble machine learning model to predict selection coefficients estimated from model simulations that span the range of malaria prevalence and treatment coverages observed globally. Specifically, we conducted model simulations that varied eleven parameters: 1) the malaria prevalence; 2) the probability of an individual seeking treatment and being effectively treated; 3) three values for the different proportions of each ACT (artemether-lumefantrine, artesunate–amodiaquine, dihydroartemisinin–piperaquine) used for front-line treatment; and 4) six values for the frequency of antimalarial resistance (K76T locus in *pfcrt*, N86Y and Y184F in *pfmdr1*, validated markers of artemisinin resistance in *pfkelch13*, copy number variation (CNV) of the *pfmdr1* gene, and CNV of the *pfpm2-3* gene(s)).

We constructed 1250 unique sets of the 11 described parameters. For all parameter combinations, ten stochastic realisations of 100,000 individuals were simulated for 40 years to reach equilibrium first before simulating the selection of *pfhrp2* deletions over the following 40 years. From each simulation we recorded the monthly resistance frequency for each of the six markers of resistance and the monthly proportion of all 28-day treatment failures. We subsequently calculated selection coefficients (the annual % change in resistance frequency or treatment failure) for each simulation repetition by linear regression of the log odds of a resistance frequency or treatment failure [10,11].

We used the generated data set to train an ensemble statistical model to predict selection coefficients based on the 11 parameters described. 25% of the simulated data sets were held back as an out-of-sample data set to be used for evaluating the performance of the trained statistical models and to test for overfitting. The remaining 75% of the simulated data was used for training three level-1 (Supplementary Figure 16) different statistical models (gradient boosted regression trees (XGBOOST), monotonic multilayer perceptron (MONMLP), and Bayesian regularised neural networks (BRNN)) to predict selection coefficients using the eleven varied transmission model parameters. Statistical model performance was evaluated based on the root mean-squared error (RMSE). Optimum model-fitting hyperparameters based on RMSE were first identified by scanning over hyperparameters for each model before fitting each model. When identifying hyperparameters for training the final model, repeated K-fold cross-validation sets were produced by splitting the training data into 5 sets of training data with the results of the cross-validation repeated 10 times to reduce any bias from the cross-validation set chosen. We calculated the performance of each final trained model by calculating the RMSE for each model when tested using the holdout data set.

To combine the predictions from each level 1 model, we used the same model fitting approach above to predict the error for each model based on the 11 parameters described. The estimated error for each level 1 model is added to the predicted selection coefficient to correct these estimates and the final predicted selection coefficient is calculated by taking the mean across each corrected estimate of selection coefficients.

Uncertainty in selection coefficients due to stochastic variation in model simulations was also estimated using a similar statistical modelling framework (75% data split, hyperparameter tuning and 20-fold cross validation). For each parameter set, we used each trained model to first predict the selection coefficient. Next, we calculate the absolute prediction error by comparing the model prediction against the selection coefficient for each stochastic realisation, before calculating the standard deviation in the error across stochastic realisations. We trained a Bayesian regularised neural network model to predict the standard deviation in error before calculating robust confidence intervals given by ±1.96 * standard deviation. To estimate the risk and future spread of antimalarial resistance in Africa, we used the weighted average ensemble model to predict selection coefficients for each resistance genotype for all admin level 1 regions based on their malaria prevalence, effective treatment coverage, use of ACTs and current resistance frequency estimated in this study. A complete schematic of this modelling pipeline is given in **Supplementary Methods Figure 3**.

The resultant selection coefficients were subsequently used to simulate the continued spread of artemisinin resistance in Africa. Given the difficulty in estimating the rate at which malaria parasites under selection spread geographically [12], we use a simple model of parasite movement to describe how resistant parasites spread between administrative regions. To simulate the spread between regions, we make the simplifying assumption that resistant parasites are exported from an admin level 1 region once the frequency of resistance is greater than 25%; when this threshold is reached, resistant parasites are seeded into neighbouring regions such that neighbouring regions reach 1% genotype frequency after one year. Once a region reaches 1% genotype frequency, the future trajectory of resistance in that region is solely determined by the selection coefficient estimated for the region for a given parameter set. Given the use of a single fixed selection coefficient for each region, this assumes that malaria prevalence and case management in each region remains constant over time. Using this approach, we simulate a range of possible timelines for the continued spread of resistance in Africa, based on using either the lower, central or upper estimate for each of the 11 parameters used to predict selection coefficients at the admin level 1 region.

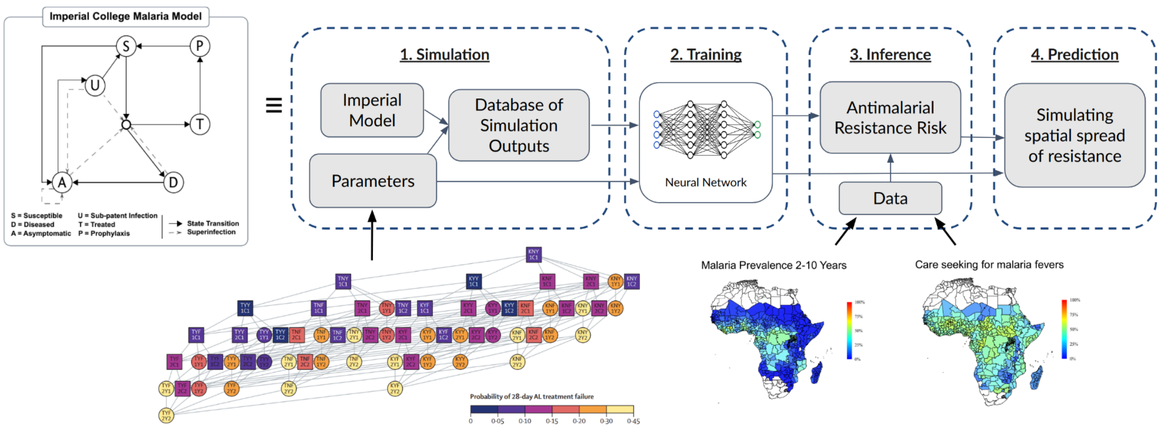


**Supplementary Methods Figure 3.** Workflow for modelling selection coefficients for antimalarial resistance in Africa and using these for prediction of future trends of resistance.

#### Supplementary Tables

**Supplementary Table 1:** Model parameters that impact the speed of selection for antimalarial resistance and data sources used to estimate these parameters. All parameters were sourced at the national level, except for malaria prevalence, effective treatment-seeking rates and resistance frequency, which were sourced at the first administrative region.

| Antimalarial resistance drivers | Impact on speed of selection for antimalarial resistance | Data Sources Used |
|---|---|---|
| Malaria Prevalence | Lower malaria prevalence will increase selection pressure by increasing the probability that individuals are infected with fewer competing strains of malaria and thus less fit resistant parasites suffer reduced interclonal competition. Additionally, lower malaria prevalence will increase the probability that an infected individual will develop a symptomatic infection (due to lower immunity at lower transmission intersities), which in turn influences the infected individual's likelihood to seek treatment. | Malaria Atlas Project maps of slide positivity ages 2-10.[13] |
| Effective Treatment Coverage for fever | Increased treatment-seeking will increase the rate at which the selective advantage conferred by antimalarial resistance is realised. Effective treatment coverage models the full treatment cascade, from seeking treatment, where treatment is sought (private vs public), whether a diagnostic test is used, and whether treatment is given for a positive, negative or no test result. | Commodities Forecast Dashboard by the Malaria Atlas Project,[14] which uses Demographic and Health Surveys (DHS), Malaria Indicator Surveys (MIS), Multiple Indicator Cluster Surveys (MICS) and AIDS Indicator Surveys (AIS) in generalized additive mixed model (GAMM) to infer treatment seeking patterns over time. |
| Non-malarial fever | Higher rates of non-malarial fevers will increase the speed of selection by increasing the rate at which asymptomatic malaria infections may seek treatment and thus allow the selective advantage to be conferred if treated. | DHS survey data with sufficiently representative fever data across all ages was used to identify an age dependent all-cause rate of fever before subtracting malaria specific fevers.[15] |
| ACT types | The use of multiple ACTs for front-line treatment will slow the speed of selection of antimalarial resistance by changing the selective pressures exerted on the parasite population.[16] | Global Fund and President's Malaria Initiative Volume of ACT sales patterns since 2019. |
| Fitness costs associated with resistance mutations. | Increased fitness costs due to antimalarial resistance will decrease the speed of selection. | Fixed multiplicative fitness cost (each additional mutation incurs additional identical costs), with an annual fitness cost of a single mutation resulting in a relative fitness of 83% is used.[1] |
| Antimalarial resistance frequency | Increased frequency of high-grade partner-drug resistance will accelerate the emergence and spread of artemisinin resistance. | Estimated in this study using WHO, WWARN & MalariaGen Pf7k resistance genotypes databases to inform spatiotemporal statistical models. |

**Supplementary Table Data: Simulated timelines for the spread of antimalarial resistance (see https://github.com/OJWatson/arms/tree/main/analysis/data-out)**

### Supplementary Figures

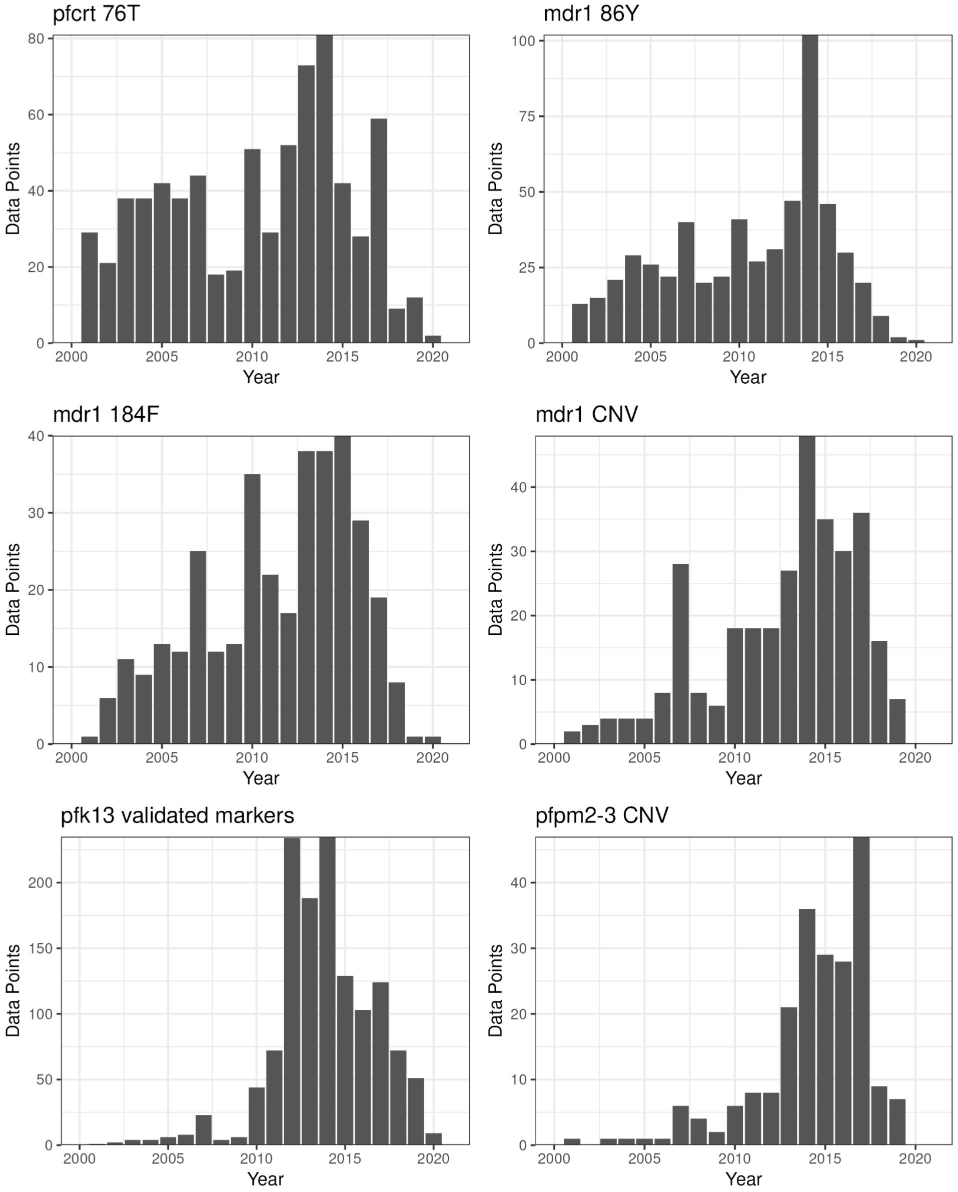


**Supplementary Figure 1.** Total number of molecular surveys in Africa for each genetic marker of resistance between 2000-2020.

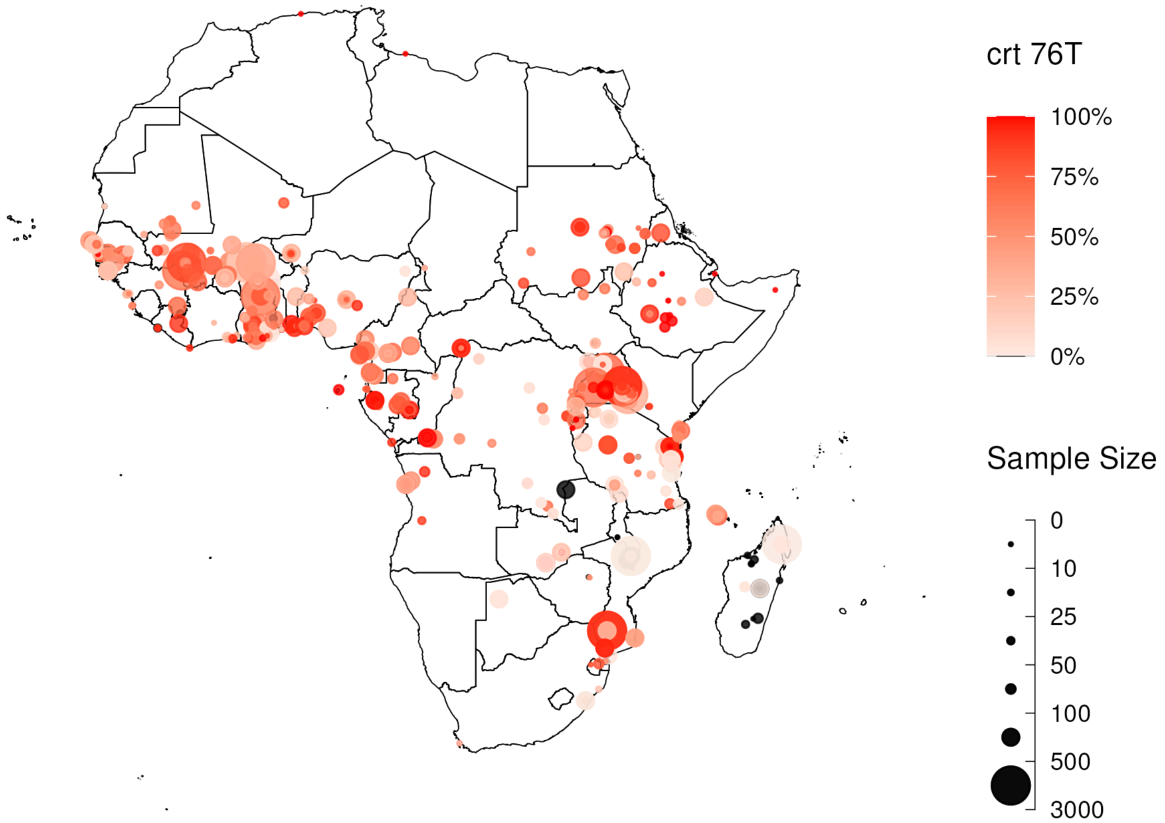


**Supplementary Figure 2.** *pfcrt 76T* mutation frequency studies 2000 - 2021 sourced from the WHO Threat Maps, WWARN and Pf7k database. Point size shows the number of samples tested for the mutation and colour indicates the frequency observed in the study, with studies with 0% frequency shown in black.

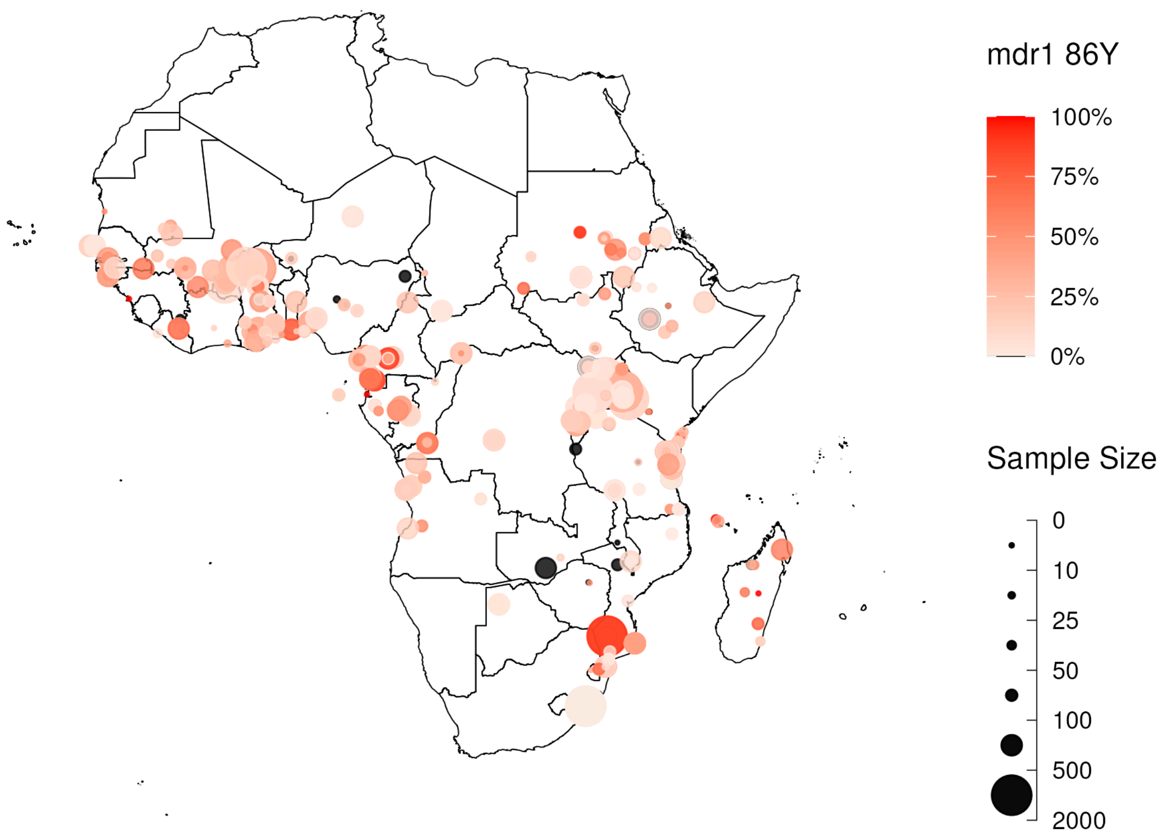


**Supplementary Figure 3.** *pfmdr1 86Y* mutation frequency studies 2000 - 2021 sourced from the WHO Threat Maps, WWARN and Pf7k database. Point size shows the number of samples tested for the mutation and colour indicates the frequency observed in the study, with studies with 0% frequency shown in black.

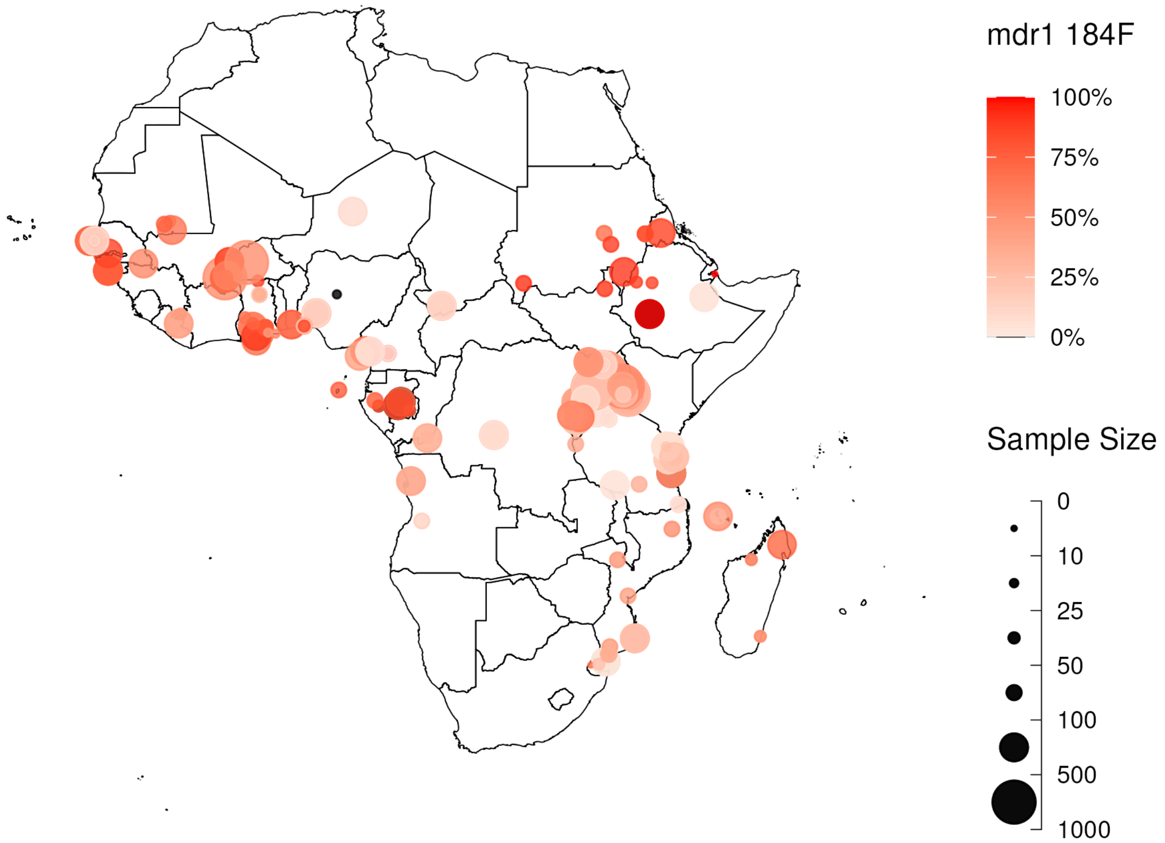


**Supplementary Figure 4.** *pfmdr1 184F* mutation frequency studies 2000 - 2021 sourced from the WHO Threat Maps, WWARN and Pf7k database. Point size shows the number of samples tested for the mutation and colour indicates the frequency observed in the study, with studies with 0% frequency shown in black.

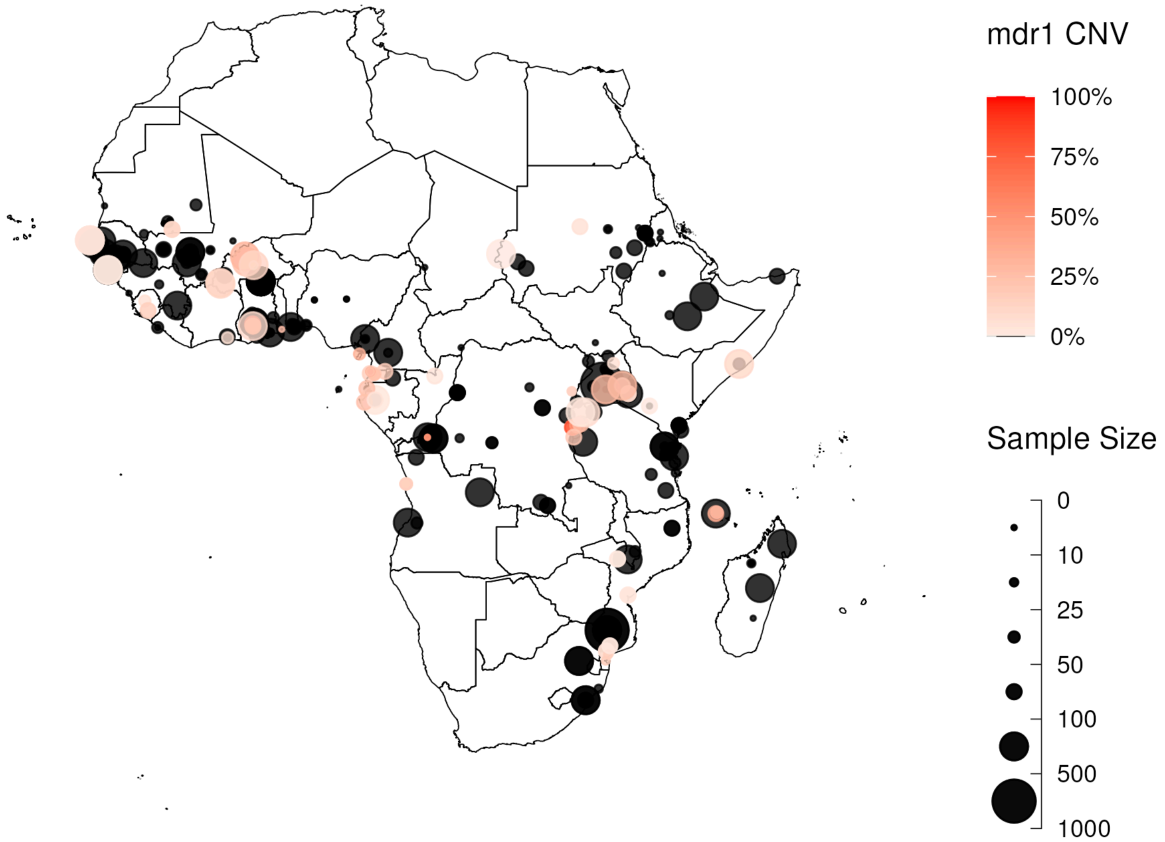


**Supplementary Figure 5.** *Pfmdr1* copy number variation frequency studies 2000 - 2021 sourced from the WHO Threat Maps, WWARN and Pf7k database. Point size shows the number of samples tested for the mutation and colour indicates the frequency observed in the study, with studies with 0% frequency shown in black.

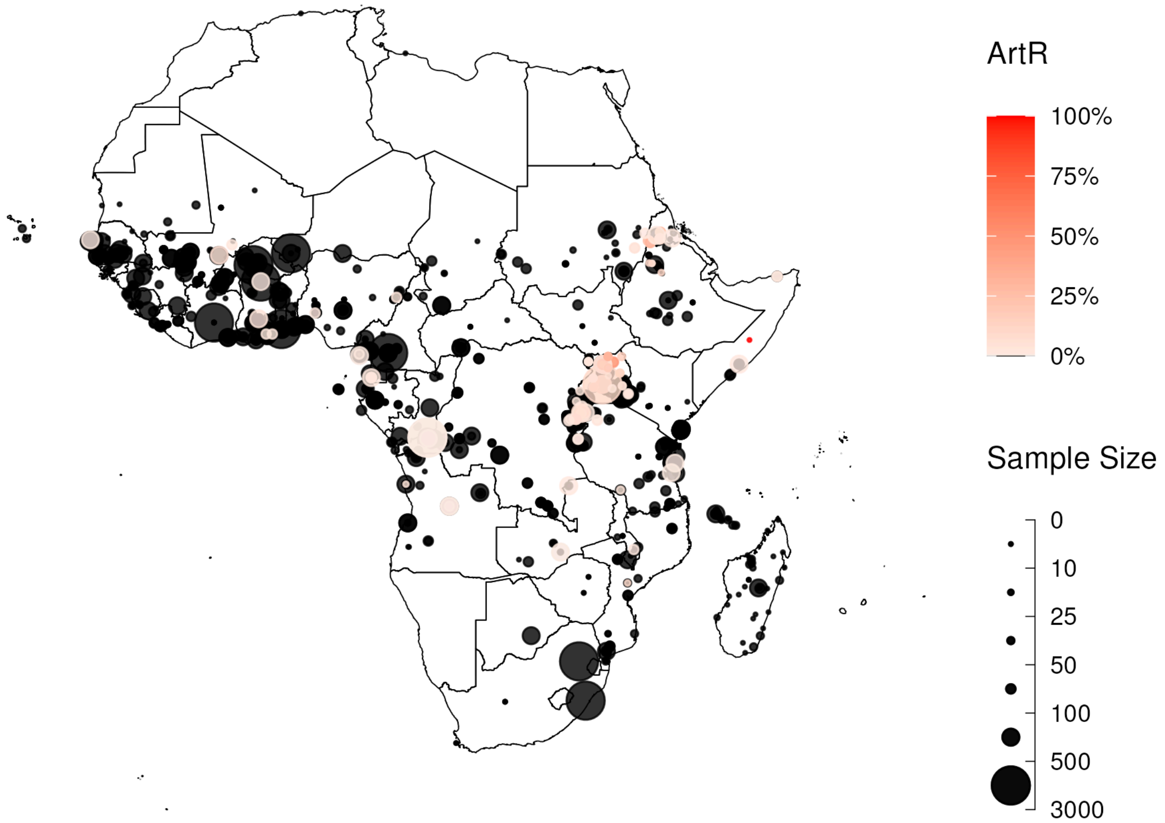


**Supplementary Figure 6.** *pfkelch13* WHO validated markers of artemisinin resistance (ART-R) frequency studies 2000 - 2021 sourced from the WHO Threat Maps, WWARN and Pf7k database. Point size shows the number of samples tested for the mutation and colour indicates the frequency observed in the study, with studies with 0% frequency shown in black.

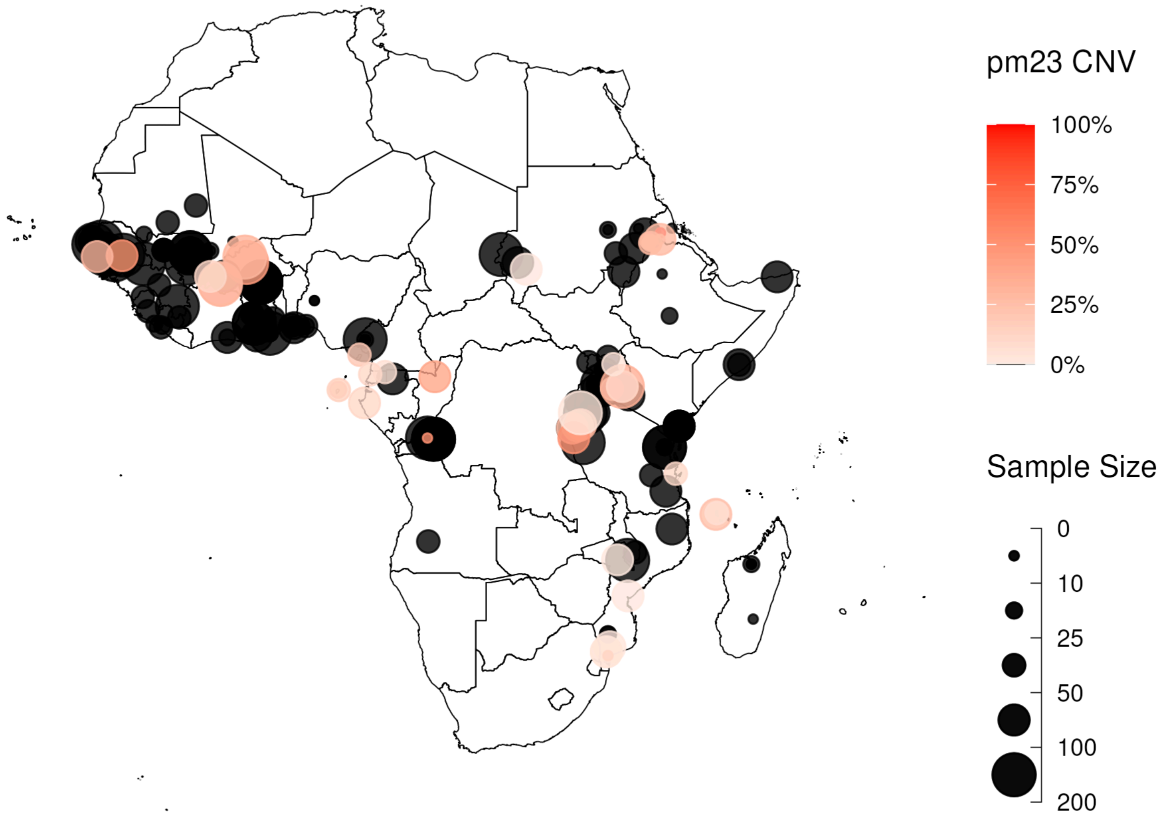


**Supplementary Figure 7.** *pfpm2-3* copy number variation frequency studies 2000 - 2021 sourced from the WHO Threat Maps, WWARN and Pf7k database. Point size shows the number of samples tested for the mutation and colour indicates the frequency observed in the study, with studies with 0% frequency shown in black.

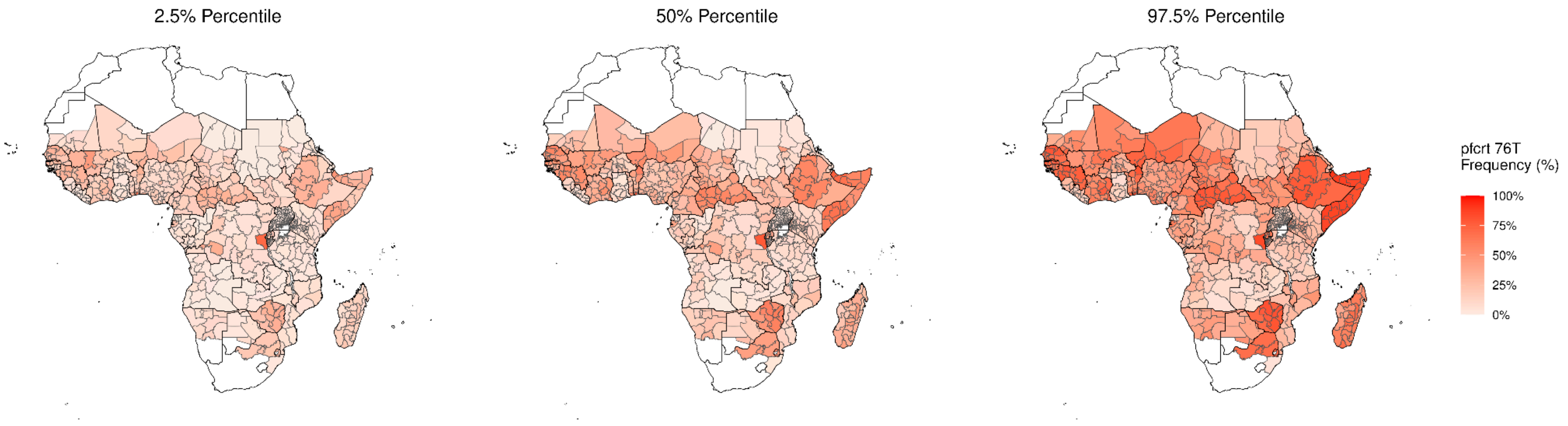


**Supplementary Figure 8.** Modelled *pfcrt 76T* mutation frequency (2.5%, 50% 97.% Credible Interval).

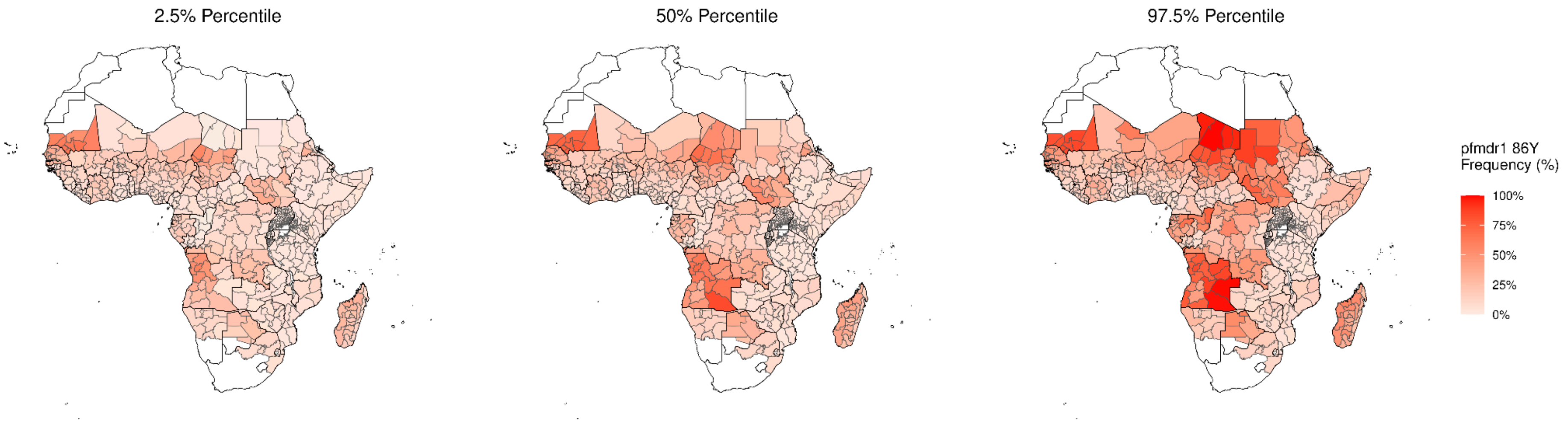


**Supplementary Figure 9.** Modelled *pfmdr1 86Y* mutation frequency (2.5%, 50%, 97.5% Credible Interval).

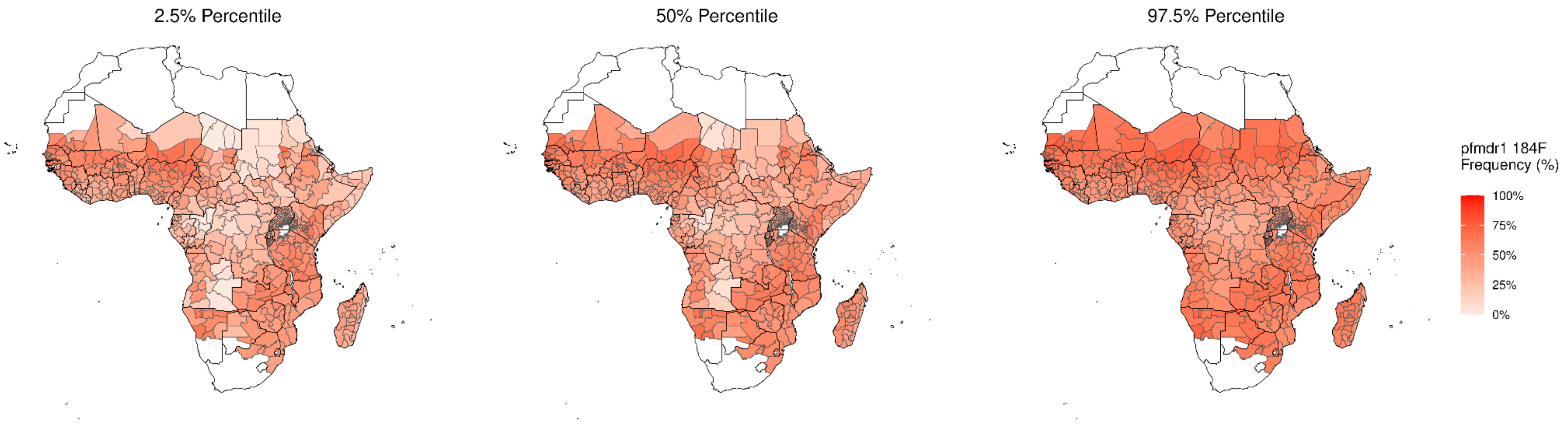


**Supplementary Figure 10.** Modelled *pfmdr1 184F* mutation frequency (2.5%, 50%, 97.5% Credible Interval).

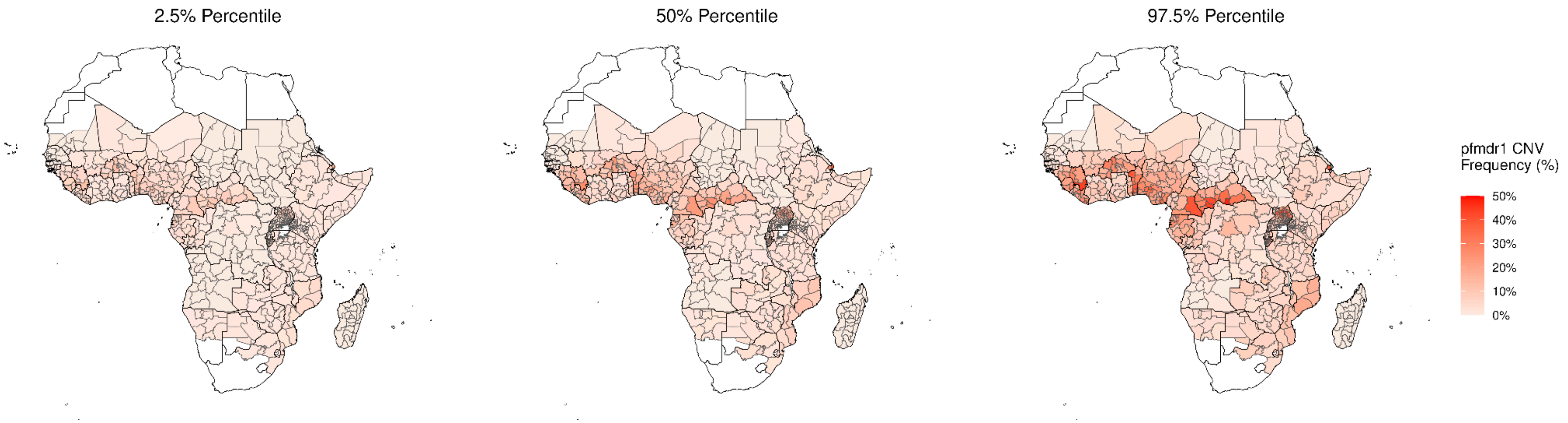


**Supplementary Figure 11.** Modelled *pfmdr1* copy number variation (CNV) mutation frequency (2.5%, 50%, 97.5% Credible Interval).

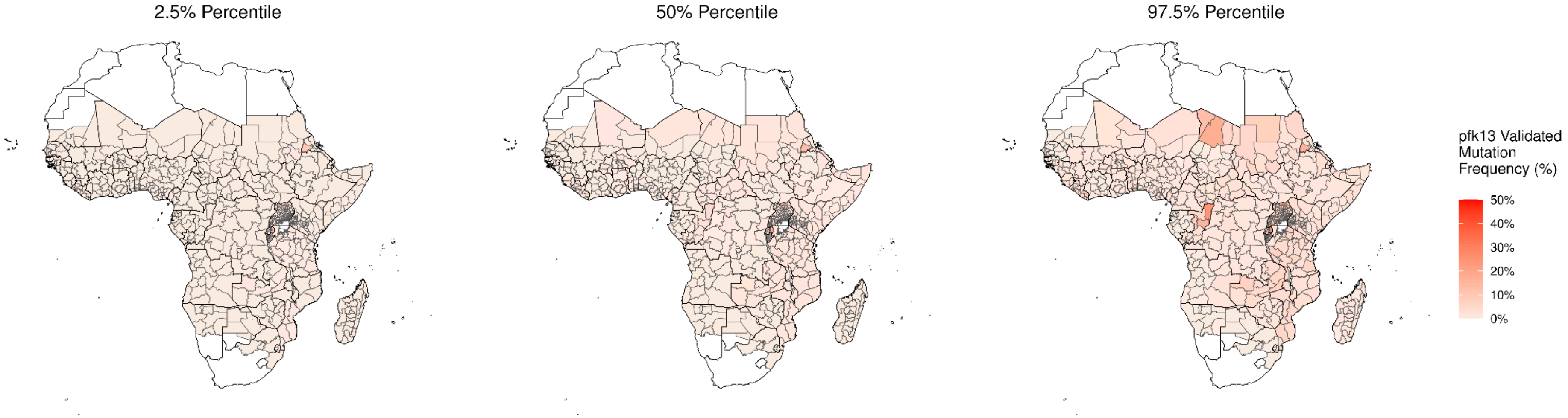


**Supplementary Figure 12.** Modelled *pfkelch13* WHO validated markers of artemisinin resistance frequency (2.5%, 50%, 97.5% Credible Interval).

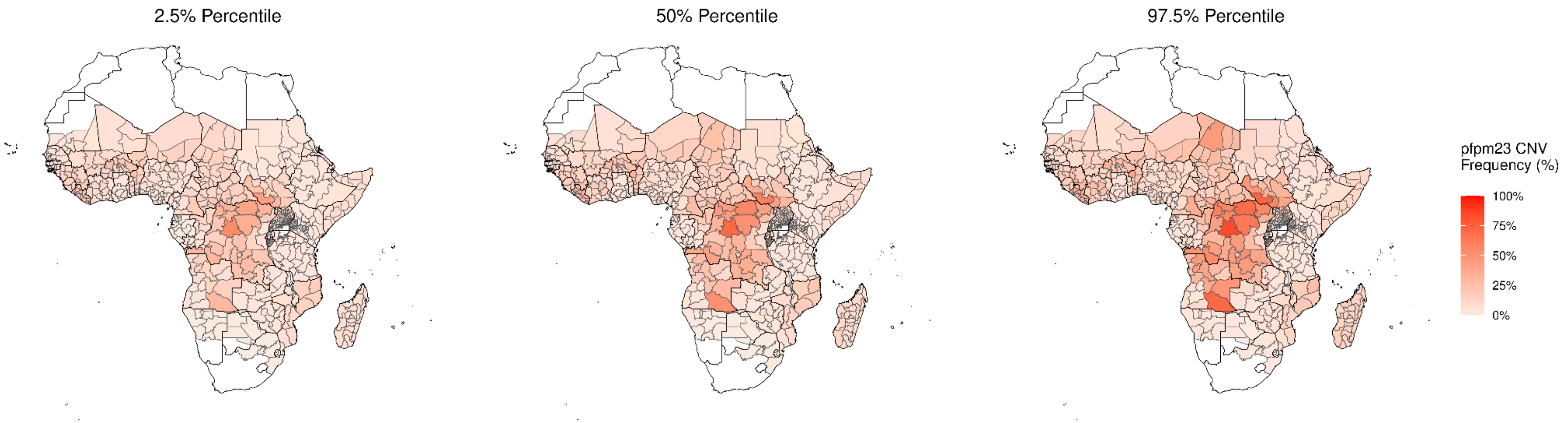


**Supplementary Figure 13.** Modelled *pfpm2-3* copy number variation (CNV) frequency (2.5%, 50%, 97.5% Credible Interval).

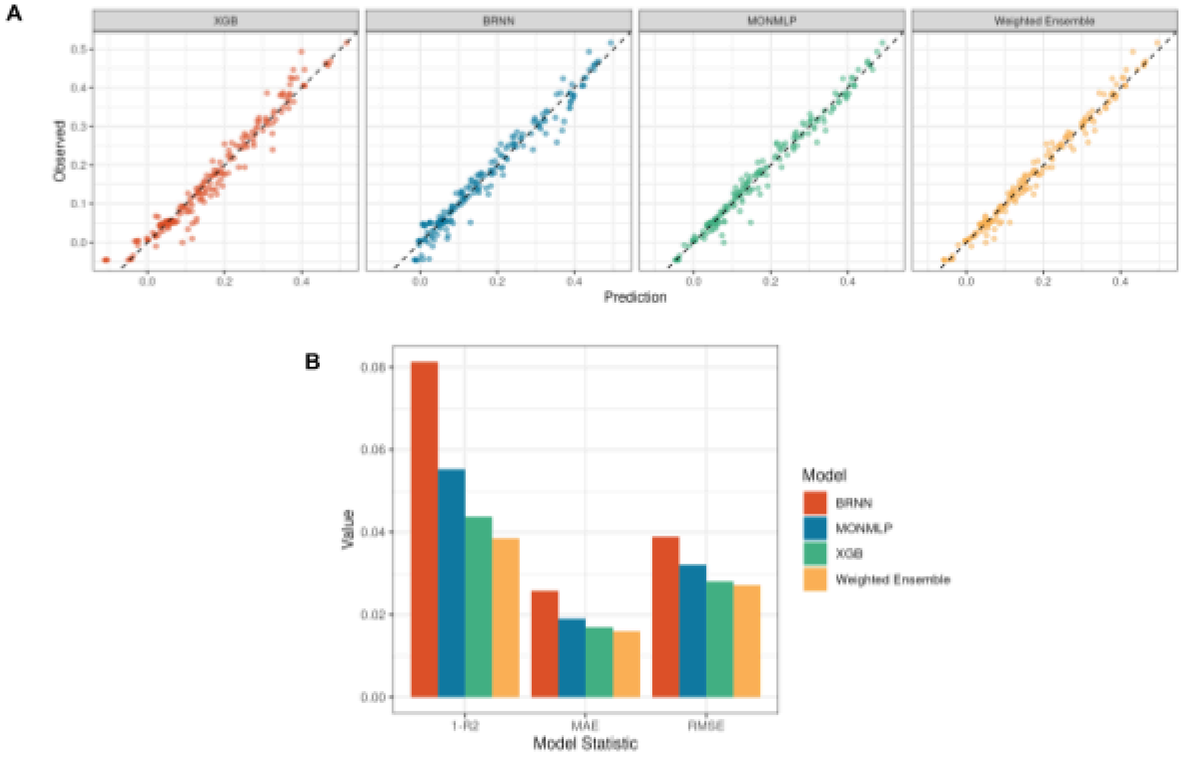


**Supplementary Figure 14.** Ensemble model fitting performance on hold-out test data. A) Model prediction selection coefficients are shown against the observed selection coefficients, with y = x trend line shown with a dashed line. B) Model performance summary statistics for each individual model alongside the weighted ensemble model.

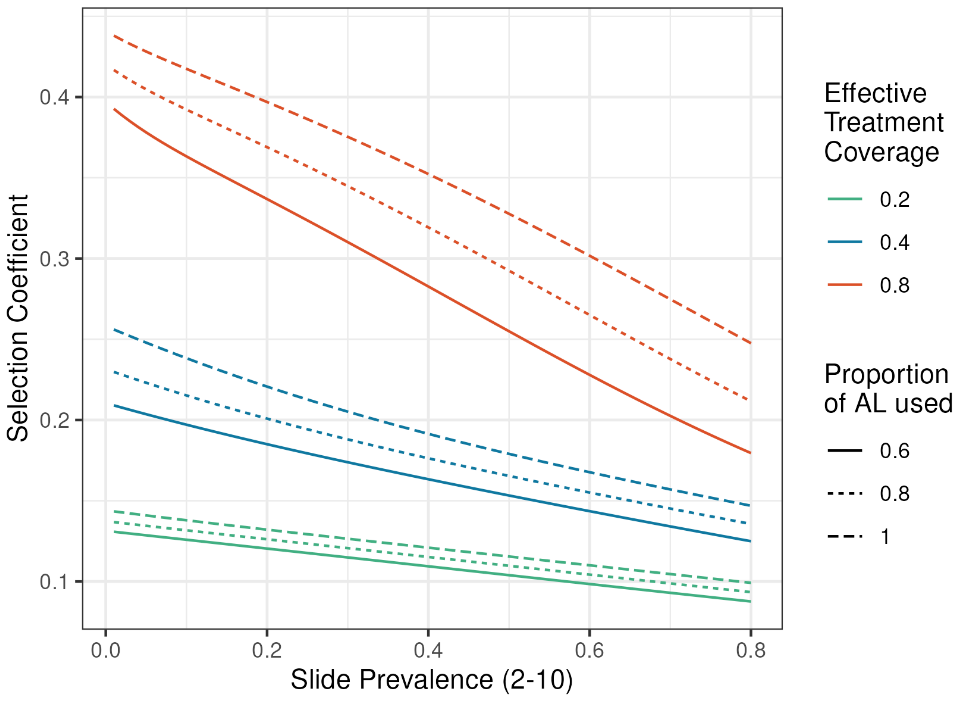


**Supplementary Figure 15.** Ensemble modelled relationship between selection coefficients for the artemisinin resistance and malaria prevalence (x-axis), treatment coverage (line colour) and proportion of artemether-lumefantrine (AL) used in front-line treatment (line type). Selection of artemisinin resistance increases as malaria prevalence decreases, the proportion of AL decreases and the effective treatment coverage increases.

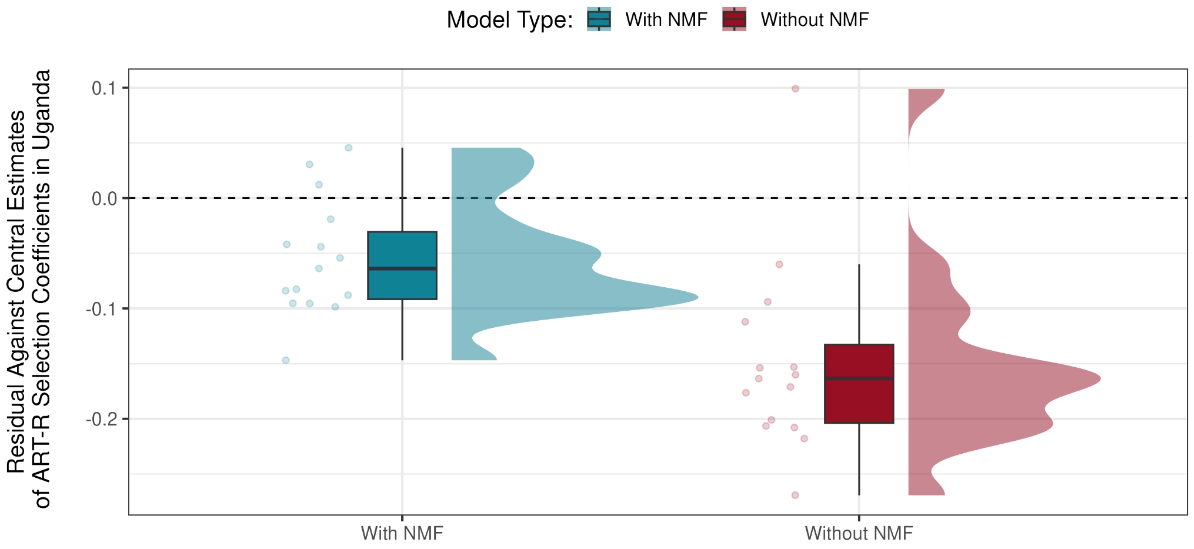


**Supplementary Figure 16.** Comparison of the agreement between simulated selection coefficients for ART-R in Uganda and the central estimates from Meier-Scherling et al. 2024 for the selection coefficients for ART-R estimated from longitudinal resistance data in Uganda. The residuals between simulated and observed selection coefficients is shown for models with non-malaria fever (NMF) in blue or without NMF in red, revealing better agreement when NMF is included.

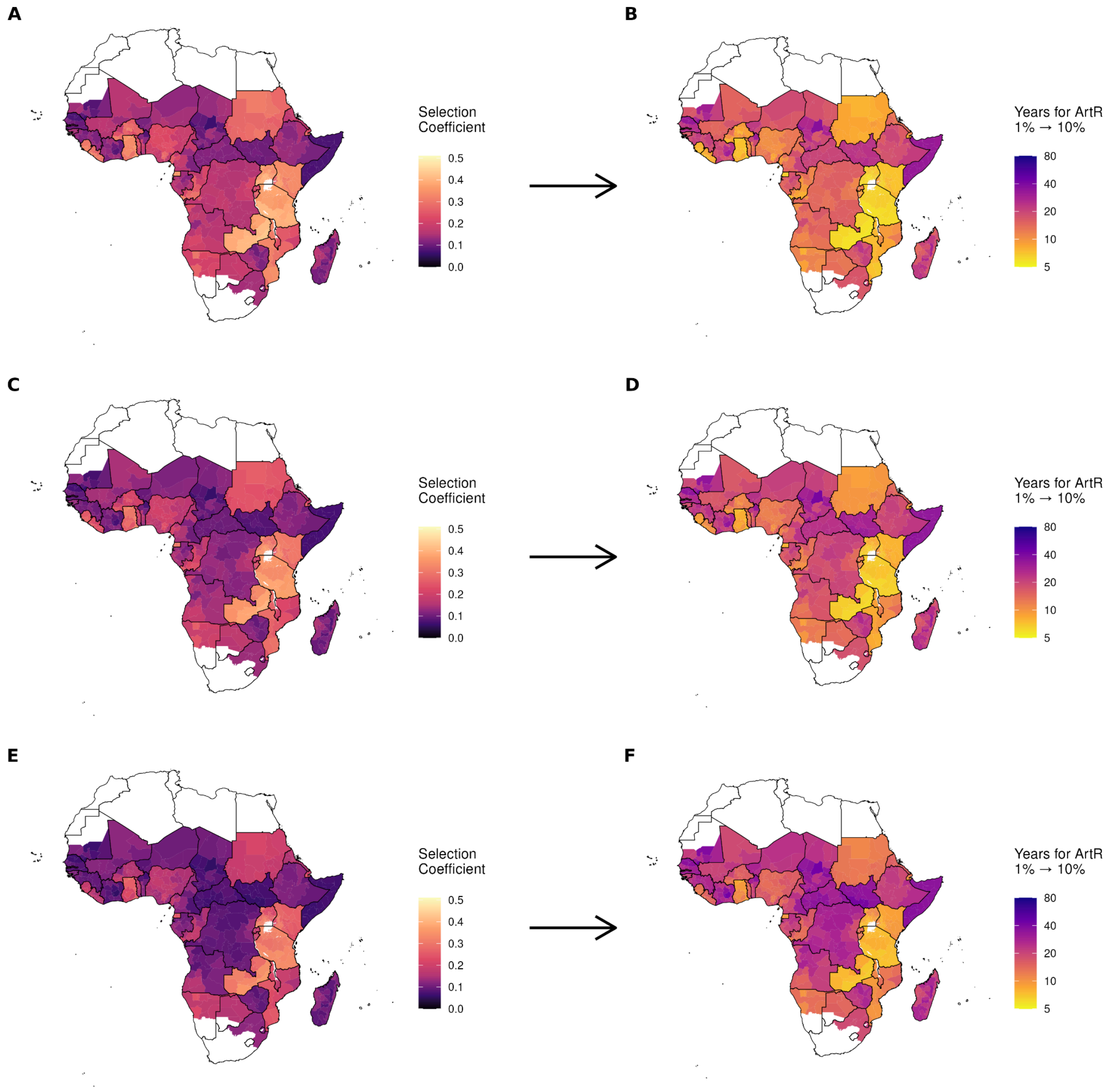


**Supplementary Figure 17.** Uncertainty in modelled selection coefficients for ART-R and equivalent times for ART-R to increase from 1% to 10%. Plots **(A)**, **(C)** and **(E)** show the modelled selection coefficient at the first-administrative region for the upper, central and lower estimates respectively. The equivalent times for ART-R to increase from 1% to 10% are shown in plots **(B)**, **(D)** and **(F)**.

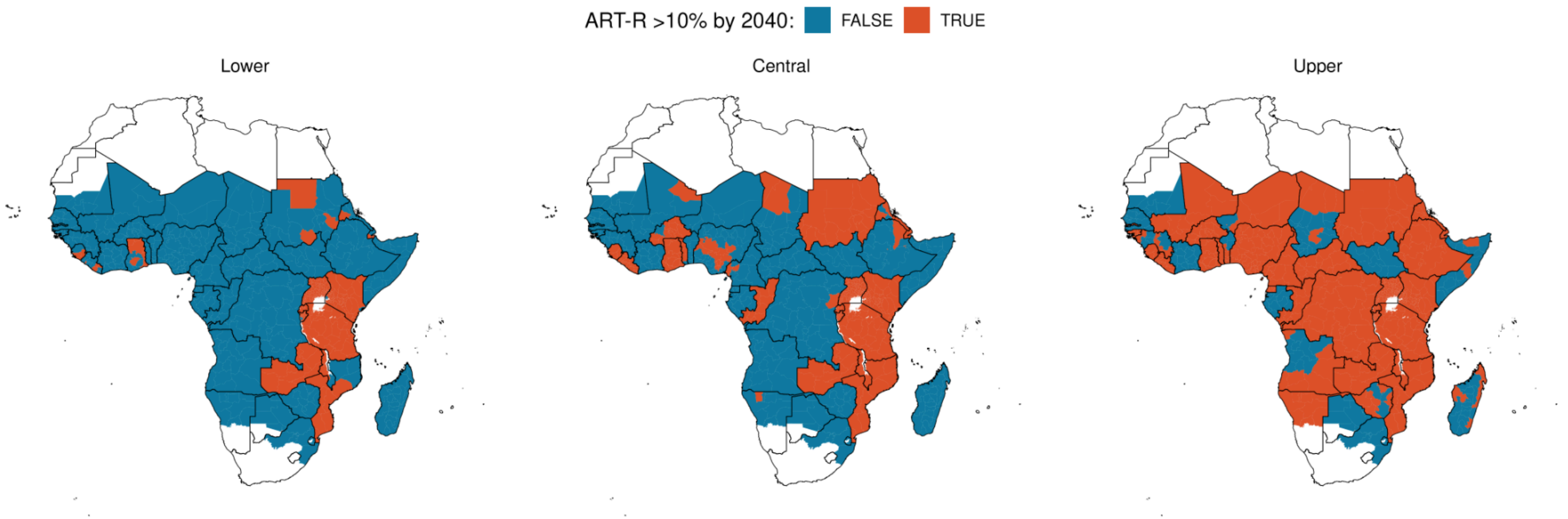


**Supplementary Figure 18.** Uncertainty in speed of ART-R spread in Africa due to uncertainty in parameter estimates of the main drivers of antimalarial resistance. Each plot shows the first-administrative regions in which ART-R is greater than 10% by 2040 for the Lower, Central and Upper estimates of parameter estimates.
